# Immune subphenotypes of tuberculosis and mortality in adults with sepsis and a high prevalence of HIV in East Africa

**DOI:** 10.64898/2025.11.29.25341077

**Authors:** Louisa Edwards, Shevin T. Jacob, Albert Majwala, Patrick Banura, Scott K. Heysell, Stellah Mpagama, Megan Null, Tania A. Thomas, Arestaricky Rimoy, Bibie Said, Edwin Nuwagira, Conrad Muzoora, Jeffrey M. Sturek, Christopher C. Moore, Eva Otoupalova

## Abstract

**Introduction:** Sepsis causes high mortality among people living with HIV in Africa, yet immune response data are limited. We identified immune subphenotypes of adults with sepsis and a high prevalence of HIV in East Africa.

**Methods:** We determined the association of serum cytokine and antibody concentrations with CD4+ T-cell and blood lactate concentration, tuberculosis, and 30-day mortality with K-means clustering, principal component analysis (PCA), and logistic regression. We validated results in a separate cohort of adults with sepsis in East Africa.

**Results:** Of 208 participants in the discovery cohort, 117 (56%) were female and 175 (84%) were living with HIV, with a mean (±SD) age of 35 (±10) years. Participants with tuberculosis had higher concentrations of G-CSF, IFN-γ, IL-1β, IL-6, IL-8, and MCP1MCAF, whereas mortality was associated with higher concentrations of G-CSF, IL-6, IL-8, IL-10, and MIP-1β, and lower concentrations of IgM antibodies against oxidation-specific epidopes (IgM^OSE^). PCA identified G-CSF, IL-5, IL-6, IL-8, and IL-13 as the main contributors to tuberculosis, and IL-4, IL-6, IL-8, IL-12, and IL-13 as the main contributors to mortality. Comprehensive biomarker and clinical and multivariable models accurately predicted tuberculosis (AUC = 0.84) and mortality (AUC = 0.78), which was replicated in the validation cohort. Cross-testing showed that the tuberculosis model delineated pathogen-specific immune activation, while the mortality model represented non-pathogen-specific immune dysregulation.

**Conclusions:** In adults with sepsis and high HIV prevalence in East Africa, tuberculosis was associated with pathogen-specific inflammation, and mortality was associated with broader immune dysregulation and diminished IgM^OSE^ antibody responses.

## INTRODUCTION

Africa has the greatest global incidence of sepsis, which causes high mortality among people living with HIV (PLWH) in whom tuberculosis is the leading cause of bloodstream infection (1, 2). Sepsis is characterized by a dysregulated immune response to infection that includes both excessive inflammation and immunosuppression through cell death, which can occur concurrently (3). While excessive inflammation can cause tissue damage, shock, and early death, immunosuppression may lead to uncontrolled infection and late mortality (4).

Advanced HIV disease is associated with cell-mediated immunosuppression and chronic perturbations of both innate and adaptive immunity, which can lead to an inadequate response to infection. Despite overall immunosuppression, PLWH may be at risk of aberrant immune activation that can lead to increased mortality (5). We found that PLWH with low CD4+ T-cell concentrations and sepsis in Uganda had a higher risk of death (6). Yet, the immune response to sepsis, including tuberculosis-related sepsis, in PLWH is not well-characterized (7).

Antibodies targeting oxidation-specific epitopes (OSEs) are evolutionarily conserved and bind to pathogens and danger-associated molecular patterns (8, 9). Anti-OSE antibodies also bind to oxidized cell membrane lipids and apoptotic cells, and can limit subsequent inflammation (8). In a tuberculosis murine model, B-1 cells, a B-cell subset known to secrete anti-OSE antibodies, attenuated mycobacterial spread (10). Antiphospholipid antibodies, similar to anti-OSE antibodies, are elevated in patients with chronic pulmonary tuberculosis and decrease with antituberculosis therapy (11, 12). Although anti-OSE antibody secreting B-1 cells are protective in murine sepsis models, they have not been evaluated in human sepsis (13–15). Given these data, anti-OSE antibodies may play a role in host defense against HIV and tuberculosis-related sepsis.

Accordingly, we investigated the association of immune subphenotypes with tuberculosis and mortality among predominantly PLWH with sepsis in Uganda. We validated our findings in a separate cohort of PLWH with sepsis from Tanzania and Uganda.

## METHODS

### Discovery cohort

We obtained blood from participants at the time of their enrollment in the PRISM-U2 fluid resuscitation study in Uganda (6). We enrolled adults aged ≥18 years admitted to Mulago National or Masaka Regional Referral Hospitals if they had clinically suspected infection, systolic blood pressure ≤100 mmHg, at least two of the following: axillary temperature >37.5°C or <35.5°C, heart rate >90 beats/min, or respiratory rate >20 breaths/min, and either a whole blood lactate concentration >2.5 mmol/L or a Karnofsky Performance Scale score ≤40. We collected blood for CD4+ T-cell and lactate concentration, standard bacterial and mycobacterial cultures, and a multiplex TaqMan Array PCR assay that included *Mycobacterium tuberculosis* (1). We tested a convenience sample of available blood specimens.

### Validation cohort

We validated our findings in blood obtained from participants at the time of their enrollment in a randomized clinical trial of early empiric anti-*Mycobacterium tuberculosis* therapy for sepsis in sub-Saharan Africa (ATLAS trial) that took place in Tanzania and Uganda at different hospitals from those included in the PRISM-U2 trial (16). We enrolled PLWH aged ≥18 years with clinically suspected infection and ≥2 modified quick sequential organ failure assessment score criteria (Glasgow coma scale score <15, respiratory rate ≥22, or systolic blood pressure ≤90 mmHg or mean arterial pressure ≤65 mmHg). We conducted the same laboratory testing as occurred in the discovery cohort and additionally tested for tuberculosis with GeneXpert on sputum and urine, and urine lipoarabinomannan antigen. We tested a subset of ATLAS blood specimens using a 5:1 discovery-to-validation ratio, with the validation cohort comprising equal numbers of participants with tuberculosis and those who died within 30 days.

### Cytokine and chemokine assays

We measured the serum concentration of 17 cytokines and chemokines: granulocyte colony-stimulating factor (G-CSF), granulocyte-macrophage colony-stimulating factor (GM-CSF), interferon-γ (IFN-γ), interleukin (IL)-1β, IL-2, IL-4, IL-5, IL-6, IL-7, IL-8, IL-10, IL-12p70, IL-13, IL-17, macrophage inflammatory protein-1β (MIP-1β), monocyte chemotactic and activating factor (MCP1MCAF), and tumor necrosis factor-α (TNF-α) using the Bio-Plex Human 17-Plex Panel (Bio-Rad Laboratories, Hercules, CA).

### Antibody assays

We diluted serum samples at a concentration of 1:10,000, 1:100,000 and 1:400,000 to measure total IgM, IgA and IgG, respectively by sandwich ELISA (Jackson ImmunoResearch Cat# 109-005-043 109-005-043, 109-005-011, and 109-005-008) as previously described (17). We established total antibody concentrations with human IgM, IgA and IgG (Thermo Scientific Cat# 39-50620-65 and 39-50550-65; Millipore Sigma Cat# I4036) standard curves. To measure concentrations of anti-OSE antibodies, anti-P1 and anti-ApoB100 IC, by sandwich ELISA, we coated plates with primary antigens P1 (Peptide2, P1 Mimitope, HSWTNSWMATFLGGGC) or capture antibody anti-ApoB100 (Millipore Sigma Cat# MAB012) and added serum samples diluted to 1:200 as previously described. We then added HRP-conjugated secondary anti-IgM, anti-IgA and anti-IgG antibodies (Jackson ImmunoResearch cat# 109-005-043 / Fisher Scientific Cat# NC0016042) in 1:40,000 dilution (Jackson ImmunoResearch Cat# 109-035-043, 109-035-011 and 109-035-190) (17). We measured absorbance at 450 nm. We normalized raw P1 and ApoB100 IC absorbances to internal controls to minimize batch-to-batch variability.

### Statistical analysis

We log-transformed all biomarker data and adjusted for potential batch effects. We used scatterplots and Pearson correlation coefficients to determine associations between biomarker concentrations, log-transformed CD4+ T-cell concentrations, and whole blood lactate. We compared biomarker concentrations between groups in the discovery and validation cohorts using Mann-Whitney U tests. To evaluate inflammatory balance, we calculated ratios of pro-inflammatory cytokines to IL-10, log-transformed them, summarized them as median (interquartile range [IQR]), and compared distributions with Mann-Whitney U tests (18). We determined associations between biomarkers and outcomes using point-biserial correlation. We performed unsupervised clustering using K-means and principal component analysis (PCA). In the PCA, we determined the biomarkers with the highest loadings. To examine relationships between tuberculosis and mortality, we disaggregated data into four groups: died with tuberculosis, died without tuberculosis, survived with tuberculosis, and survived without tuberculosis.

For multivariable logistic regression models, we selected biomarkers based on statistical significance in univariable analyses of the discovery cohort. In each cohort, we built separate multivariable models to predict tuberculosis or 30-day mortality: 1) biomarker models, 2) comprehensive models that included biomarkers and clinical variables (lactate and HIV serostatus), and 3) parsimonious clinical models using the Universal Vital Assessment (UVA) mortality risk score and the single biomarker with the greatest univariable association with the outcome of interest (19). To adjust for potential confounding, we included age and sex as covariates in all models. We evaluated model performance in each cohort using the area under the receiver operating characteristic curve (AUC) with 95% confidence intervals (CIs) estimated from 1,000 bootstrapped resamples. We evaluated model calibration by plotting predicted probabilities against observed event frequencies. We tested the multivariable models on disaggregated data to evaluate the separate effects of tuberculosis and mortality in the discovery cohort. We classified participants who died and those who survived into three infection groups: 1) confirmed tuberculosis, 2) no confirmed tuberculosis but infected with a non-tuberculosis pathogen, or 3) no identified pathogen and then conducted multinomial logistic regression with participants who survived and had no identified pathogen as the referent group.

All p-values were two-tailed. We defined statistical significance as p<0.05 after correcting for multiple comparisons using the Benjamini-Hochberg procedure (20). We conducted all analyses using R software (R Core Team [2024]. R: A language and environment for statistical computing. R Foundation for Statistical Computing, Vienna, Austria. URL http://www.R-project.org/).

### Ethical considerations

We obtained approval for the PRISM-U2 trial from the Uganda National Council of Science and Technology (UNCST HS#419) and the University of Virginia (HSR# 13393). We obtained approval for the ATLAS trial from the Tanzania Medicines and Medical Devices Authority (TMDA-WEB0021/CTR/0008/03) and National Institute of Medical Research (NIMR/HQ/R.8a/Vol.IX/3664), Uganda National Council of Science and Technology (UNCST HS1272ES) and the University of Virginia (HSR#200253) and registered at clinicaltrials.gov (NCT04618198). Participants or their next of kin provided informed consent prior to enrollment.

## RESULTS

### Discovery and validation cohort characteristics

We sampled 208 of 426 (49%) PRISM-U2 trial participants for the discovery cohort; 117 (56%) were female, the mean (± standard deviation [SD]) age was 35 (±10) years, the median (IQR) lactate concentration was 3.7 (2.9-4.7) mmol/L and 64 (31%) had confirmed tuberculosis **(Table 1)**. There were 175 (84%) PLWH with a median (IQR) CD4+ T-cell concentration of 37 (11–111) cells/µL. There were 74 (38%) participants who died by day-30. We sampled 40 of 395 (10%) ATLAS trial participants for the validation cohort, 23 (58%) were female, the mean (±SD) age was 43 (±13) years, the median (IQR) lactate concentration was 2.5 (1.6-3.1) mmol/L and 20 (50%) had confirmed tuberculosis **(Table 1)**. All were PLWH with a median (IQR) CD4+ T-cell concentration of 70 (25–244) cells/µL. There were 20 (50%) participants who died by day-30 **(Figure S1)**.

**Table 1.** Characteristics and outcomes of the PRISM-U2 discovery cohort and the ATLAS trial validation cohort.

|  | PRISM U-2 Discovery Cohort |  |  |  |  | ATLAS Validation Cohort |  |  |  |  |
| --- | --- | --- | --- | --- | --- | --- | --- | --- | --- | --- |
|  | All<br>N=194 | Died /<br>Confirmed TB<br>N=35 | Died /<br>No TB<br>N=39 | Survived /<br>Confirmed TB<br>N=27 | Survived /<br>No TB<br>N=93 | All<br>N=40 | Died /<br>Confirmed TB<br>N=10 | Died /<br>No TB<br>N=10 | Survived /<br>Confirmed TB<br>N=10 | Survived /<br>No TB<br>N=10 |
| Age, years (mean ±SD) | 35 (10.0) | 36 (10.0) | 34 (8.0) | 36 (8.0) | 35 (12.0) | 43 (13.0) | 47 (14) | 39 (13.5) | 39 (10.0) | 47 (13) |
| Female (%) | 117 (56.2) | 14 (40.0) | 27 (69.2) | 9 (33.3) | 60 (64.5) | 23 (57.5) | 5 (50.0) | 7 (70.0) | 5 (50.0) | 5 (50.0) |
| Living with HIV (%) | 175 (84.1) | 32 (91.4) | 37 (94.9) | 26 (96.3) | 71 (76.3) | 40 (100.0) | 10 (100) | 10 (100) | 10 (100) | 10 (100) |
| Confirmed tuberculosis (%) | 64 (30.8) | 35 (100) | 0 (0) | 27 (100) | 0 (0) | 20 (50.0) | 10 (100) | 0 (0) | 10 (100) | 0 (0) |
| Lactate, mmol/L (mean ±SD) | 4.1 (2.0) | 5.0 (2.8) | 4.4 (2.4) | 3.8 (1.3) | 3.7 (1.7) | 2.5 (1.1) | 2.6 (1.1) | 2.1 (1.0) | 2.9 (1.5) | 2.4 (1.0) |
| UVA score, n (mean ±SD) | 4.5 (1.7) | 5.1 (1.8) | 4.6 (1.7) | 4.5 (1.6) | 4.4 (1.7) | 6.2 (2.3) | 6.7 (2.5) | 6.6 (1.9) | 6.1 (2.6) | 5.2 (2.2) |
| Died by 30 days (%) | 74 (38.1) | 35 (100) | 39 (100) | 0 (0) | 0 (0) | 20 (50.0) | 10 (100) | 10 (100) | 0 (0) | 0 (0) |

### Biomarkers in the discovery cohort

#### Correlation, K-means clustering, and principal component analysis

CD4+ T-cell and lactate concentrations had weak correlations with biomarker concentrations **(Figures S2-5)**. We observed positive pairwise correlations among G-CSF, GM-CSF, IFN-γ, IL-4, IL-5, IL-12, IL-13, IL-17, MCP1MCAF, and TNF-α **(Figure S6)**. Negative pairwise correlations occurred between anti-ApoB100 IC IgA and MCP1MCAF, and anti-ApoB100 IC IgG and MIP-1β. K-means identified two clusters: 1) G-CSF, IFN-γ, IL-1β, IL-4, IL-5, IL-7, IL-12, IL-13, and TNF-α, and 2) G-CSF, IL-6, IL-8, IL-17, MCP1MCAF, and MIP-1β **(Figure S7)**. PCA identified G-CSF, IL-6, IL-8, IL-13, and MIP-1β as the top-loading biomarkers **(Table S1; Figure 1)**.

**Figure 1.**
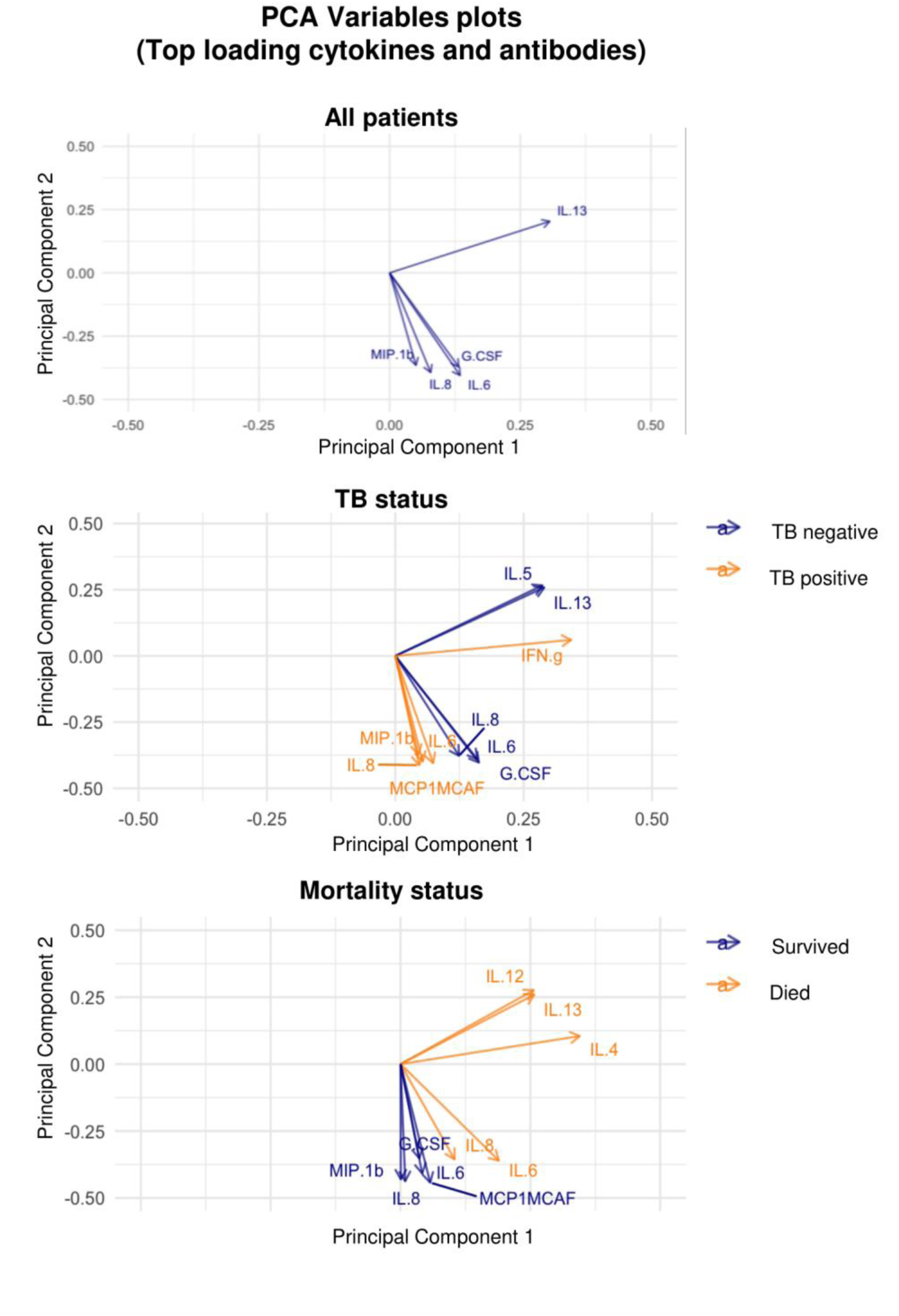
Principal component analysis of the five highest-loading cytokines and antibodies among adults with sepsis in Uganda included in. **A)** the entire discovery cohort, **B)** stratified by tuberculosis status, and **C)** stratified by mortality.

### Biomarkers and tuberculosis

#### Univariable analyses

In the discovery cohort, participants with tuberculosis had significantly higher median concentrations of G-CSF, IFN-g, IL-1β, IL-6, IL-8, and MCP1MCAF compared to participants without tuberculosis **(Figure 2)**. Antibodies had overlapping distributions between groups **(Figure 2)**. Compared to participants without tuberculosis, distribution of cytokine-to-IL-10 ratios differed for IL-1β (significantly higher) and IL-6 (significantly lower) among participants with tuberculosis **(Figure S8)**. In the univariable analysis, IL-1β, IL-8, and MCP1MCAF were associated with tuberculosis (p<0.05) **(Table S2, Figure 3, and Figures S9 and S10)**. After disaggregation, tuberculosis was associated with increased IL-1β, IL-8, and total IgM, whether or not the participant died **(Figures 2 and 3)**. With the exception of IL-7, the ratios of cytokines to IL-10 were similarly pro-inflammatory **(Figure S11)** and were replicated in the validation cohort **(Figures S12 and S13)**.

**Figure 2.**
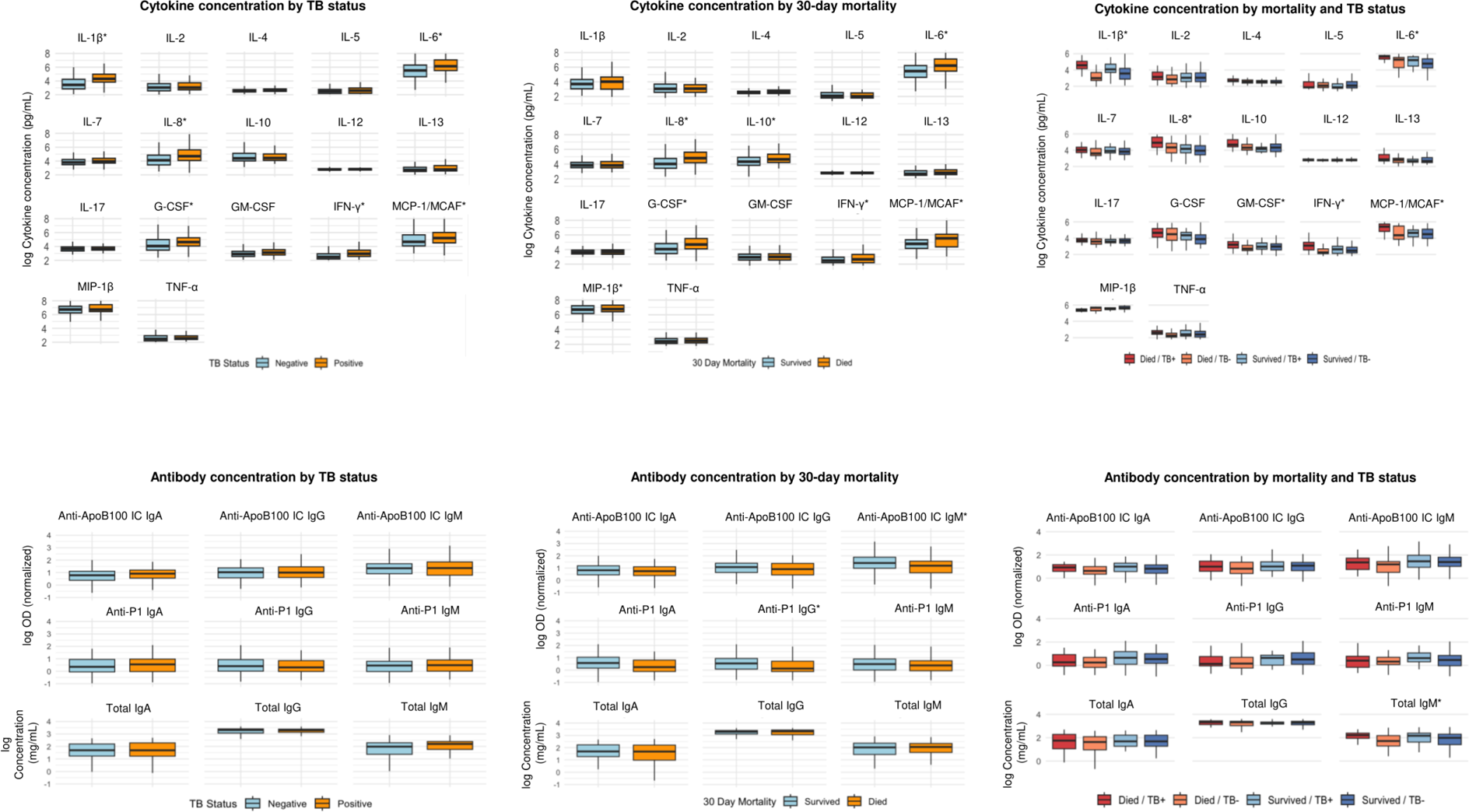
Box-and-whisker plots showing the median, interquartile range, and overall distribution of log-transformed biomarker concentrations among adults with sepsis in Uganda included in the discovery cohort.

**Figure 3.**
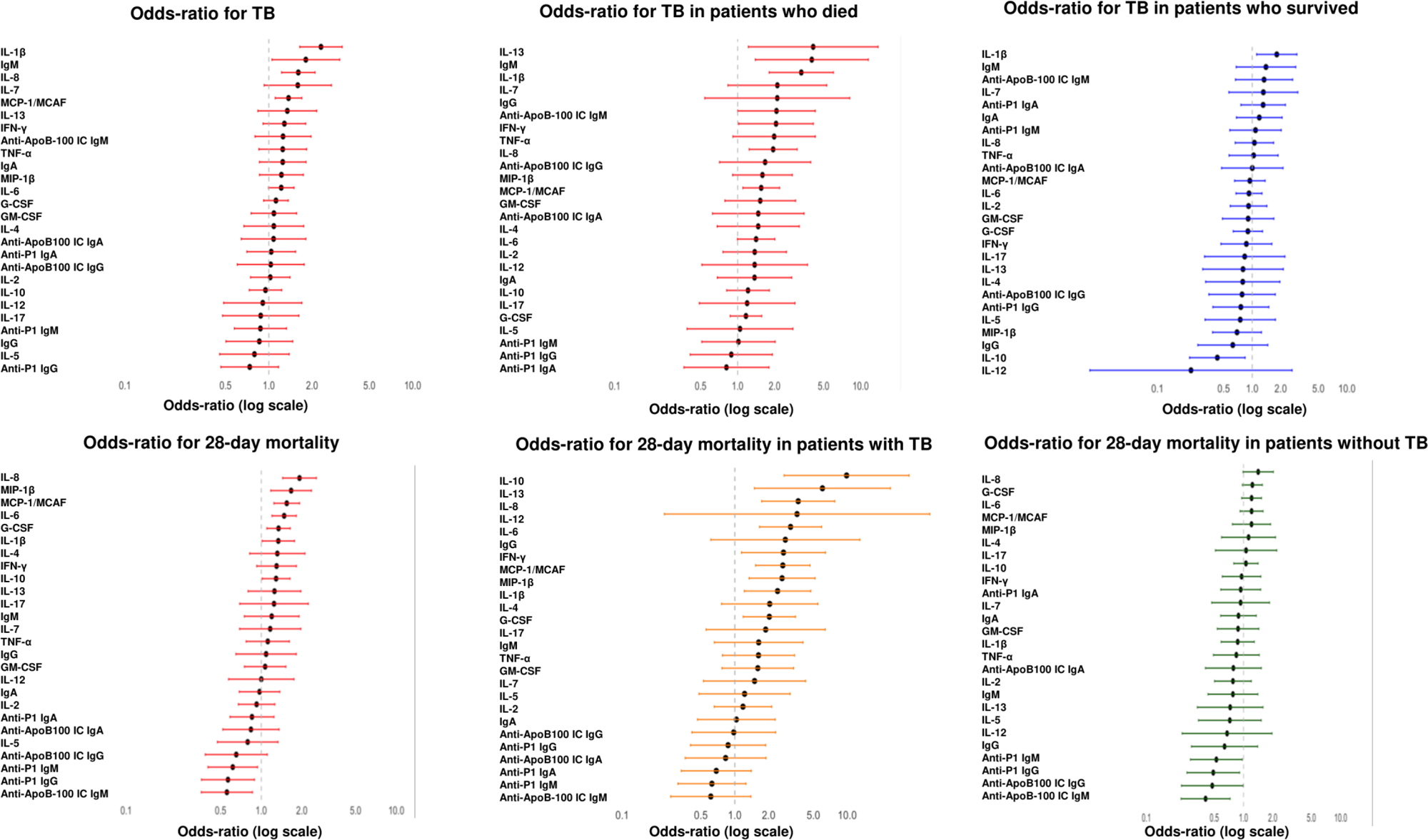
Forest plots showing odds ratios and 95% confidence intervals for associations between biomarkers and clinical outcomes among adults with sepsis in Uganda included in the discovery cohort.

#### Correlation, K-means clustering, and principal component analysis

The pattern of biomarker correlations in participants with tuberculosis was consistent with that of the entire cohort **(Figure S14)**. Negative pairwise correlations included anti-P1 IgM with IgA, anti-ApoB100 IC IgA with IL-8, and total IgA with IL-7. Clusters were consistent across tuberculosis status with the most distinct separation between clusters in participants without tuberculosis **(Figure S7)**. After disaggregation, clusters were similar between participants who died and those who survived within each tuberculosis status group. Among participants with tuberculosis, we identified a subgroup with elevated GM-CSF, IFN-γ, IL-2, IL-4, and TNF-α regardless of mortality status **(Figure S7)**. PCA identified G-CSF, IL-5, IL-6, IL-8, and IL-13 as most associated with tuberculosis, and IFN-γ, IL-6, IL-8, MCP1MCAF, and MIP-1β as most associated with participants without tuberculosis **(Table S1; Figure 1)**.

#### Multivariable logistic regression models of tuberculosis

A multivariable biomarker model that included IL-1β and IL-8 predicted tuberculosis with an AUC of 0.80 (95% CI 0.73-0.86) **(Table 2 and Figure 4)**. This biomarker model plus total IgM predicted tuberculosis with an AUC of 0.81 (95% CI 0.74-0.87). A comprehensive biomarker and clinical multivariable model that added HIV serostatus and lactate to the prior model predicted tuberculosis with an AUC of 0.84 (95% CI 0.77-0.89). The addition of IL-1β and lactate to UVA score improved the discrimination of tuberculosis from 0.59 (95% CI 0.50-0.68) to 0.68 (95% CI 0.58-0.81). We replicated these results in the validation cohort **(Table 2)**. The comprehensive model had the best calibration, while the biomarker models that included cytokines and antibodies overpredicted when the observed tuberculosis rate was moderate-to-high **(Figure 4)**. After disaggregation, the tuberculosis models performed best among participants who died **(Table S4)**.

**Figure 4.**
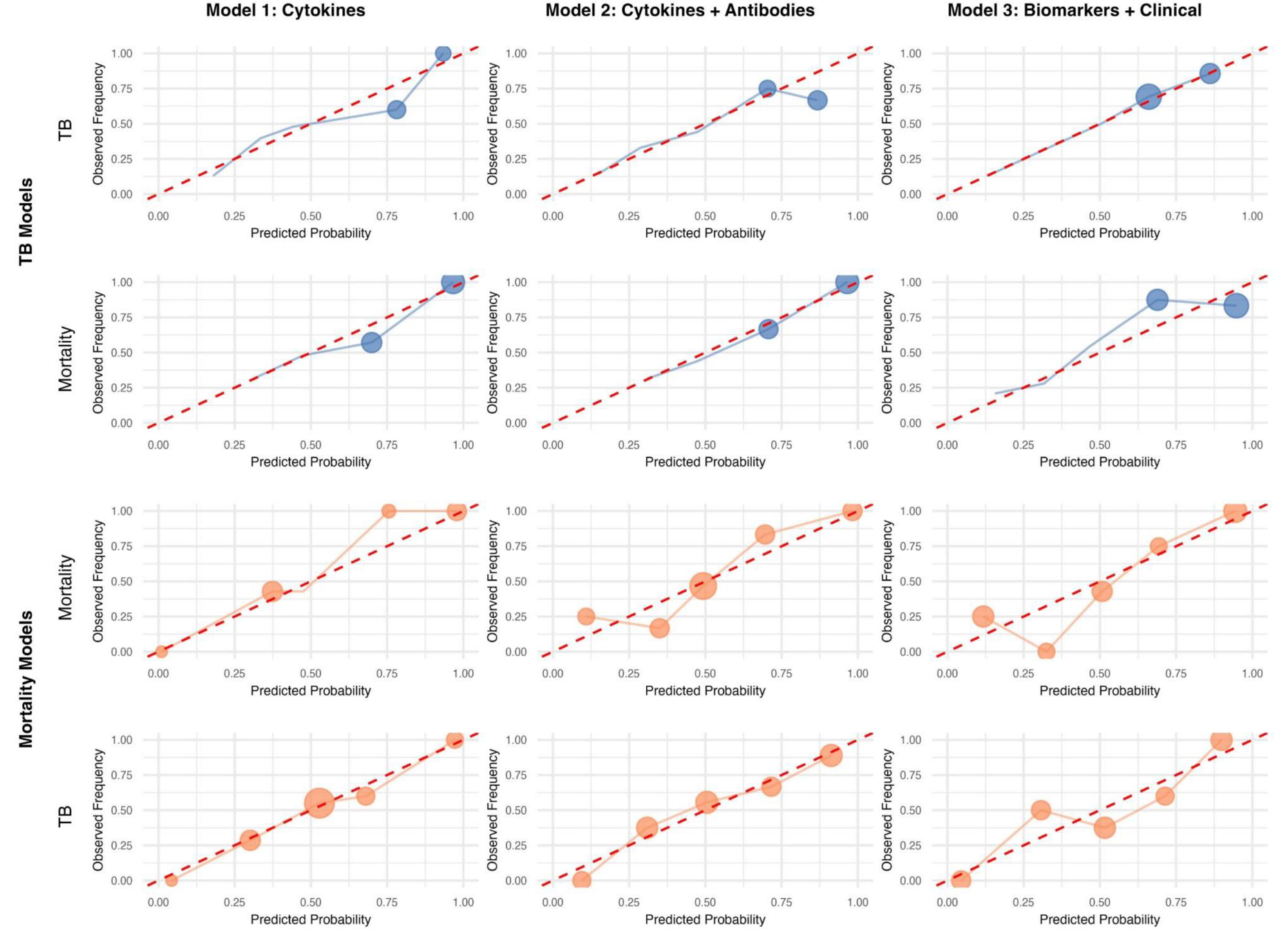
Calibration curves for multivariable logistic regression models predicting tuberculosis and mortality among adults with sepsis in Uganda included in the discovery cohort. Both the tuberculosis and mortality models are applied to participants stratified by tuberculosis status (first and fourth rows of panels) and mortality status (second and third rows of panels). Model 1 includes only cytokines; Model 2 includes cytokines and antibodies; and Model 3 includes cytokines, antibodies, HIV status, and lactate concentration. Circles on the calibration curves represent bins of patients grouped by their predicted probabilities of the outcome, while the size of the circles represents the number of patients in each bin. Values above the diagonal dashed line indicate underprediction, whereas points below the line indicate overprediction of risk.

**Table 2.** Multivariable prediction models of tuberculosis and 30-day mortality among adults with sepsis in East Africa.

|  | Discovery cohort |  | Validation cohort |  |
| --- | --- | --- | --- | --- |
|  | AUC (95% CI) | P-value | AUC (95% CI) | P-value |
| **Biomarker models of tuberculosis** |  |  |  |  |
| Age + Sex + IL-1β + IL-8 | 0.80<br>(0.73–0.86) | Referent | 0.86<br>(0.61–1.00) | Referent |
| Age + Sex + IL-1β + IL-8 + Total IgM | 0.81<br>(0.74–0.87) | 0.33 | 0.91<br>(0.67–1.00) | 0.18 |
| Age + Sex + IL-1β + IL-8 + Total IgM + Lactate + HIV | 0.84<br>(0.77–0.89) | 0.40 | 0.97<br>(0.76–1.00) | 0.37 |
| **Clinical models of tuberculosis** |  |  |  |  |
| UVA score | 0.59<br>(0.50–0.68) | Referent | 0.58<br>(0.44–0.75) | Referent |
| UVA score + Lactate | 0.61<br>(0.50–0.69) | 0.83 | 0.68<br>(0.45–0.86) | 0.46 |
| UVA score + Lactate + IL-8 | 0.65<br>(0.56–0.74) | 0.07 | 0.70<br>(0.43–0.90) | 0.40 |
| UVA score + Lactate + IL-1β | 0.68<br>(0.58–0.81) | 0.02 | 0.84<br>(0.55–1.00) | 0.07 |
| **Biomarker models of mortality** |  |  |  |  |
| Age + Sex + IL-6 + IL-8 + IL-10 + MIP-1β | 0.72<br>(0.64–0.80) | Referent | 0.83<br>(0.66–0.96) | Referent |
| Age + Sex + IL-6 + IL-8 + IL-10 + MIP-1β + anti-P1 IgM + anti-ApoB100 IC IgM | 0.74<br>(0.67–0.81) | 0.22 | 0.85<br>(0.70–0.96) | 0.44 |
| Age + Sex + IL-6 + IL-8 + IL-10 + MIP-1β + anti-P1 IgM + anti-ApoB100 IC IgM + Lactate + HIV | 0.78<br>(0.70–0.84) | 0.50 | 0.88<br>(0.75–0.97) | 0.13 |
| **Clinical models of mortality** |  |  |  |  |
| UVA score | 0.57<br>(0.49–0.65) | Referent | 0.62<br>(0.43–0.80) | Referent |
| UVA score + Lactate | 0.62<br>(0.53–0.71) | 0.25 | 0.65<br>(0.42–0.83) | 0.34 |
| UVA score + Lactate + IL-10 | 0.63<br>(0.53–0.72) | 0.23 | 0.82<br>(0.67–0.94) | 0.29 |
| UVA score + Lactate + IL-8 | 0.71<br>(0.63–0.78) | 0.002 | 0.68<br>(0.43–0.85) | 0.22 |
| **Comprehensive tuberculosis model predicting mortality** |  |  |  |  |
| Age + Sex + IL-1β + IL-8 + Total IgM + Lactate + HIV | 0.77<br>(0.69–0.84) | 0.06* | 1.00<br>(0.92–1.00) | 0.33* |
| **Comprehensive mortality model predicting tuberculosis** |  |  |  |  |
| Age + Sex + IL-6 + IL-8 + IL-10 + MIP-1β + anti-P1 IgM + anti-ApoB100 IC IgM + Lactate + HIV | 0.81<br>(0.74–0.87) | 0.66** | 0.90<br>(0.77–0.97) | 0.90** |

AUC, area under the receiver operating characteristic curve; CI, confidence interval; IL, interleukin; MIP, Macrophage Inflammatory Protein; UVA, Universal Vital Assessment mortality risk score. P-values were calculated using DeLong tests comparing each AUC with the referent model AUC; \*P-values reflect DeLong comparisons with the same model's AUC for tuberculosis prediction; \*\*P-values reflect DeLong comparisons with the same model's AUC for mortality prediction.

### Biomarkers and mortality

#### Univariable analyses

In the discovery cohort, participants who died had significantly higher median concentrations of G-CSF, IL-6, IL-8, IL-10, MCP1MCAF, and MIP-1β than participants who survived **(Figure 2)**, while participants who survived had a significantly higher median serum concentration of anti-ApoB IgM and anti-P1 IgG **(Figure 2)**. Compared to participants who survived, the distribution of cytokine-to-IL-10 ratios for IL-2, IL-6, IL-7, IL-12, IL-17, and TNF-α differed significantly from those participants who died **(Figure S15)**. All ratios except IL-6:IL-10 were higher in participants who survived. In the univariable analysis, G-CSF, IL-6, IL-8, MCP1MCAF, and MIP-1β were associated with increased mortality (p<0.05), while anti-ApoB IgM and anti-P1 IgG were associated with decreased mortality **(Table S3, Figure 3, and Figures S9 and S10)**. After disaggregation, death with tuberculosis was associated with the greatest concentration of pro-inflammatory cytokines and survival without tuberculosis was associated with the lowest concentration of pro-inflammatory cytokines **(Figure 2)**. Survival with tuberculosis was associated with greater inflammation compared to death without tuberculosis, which was most notable with GM-CSF, IL-1β, IL-6, and MCP1MCAF. The ratios of cytokines to IL-10 were similarly pro-inflammatory **(Figure S11)** and were replicated in the validation cohort **(Figures S12 and S13)**.

#### Correlation, K-means clustering, and principal component analysis

The pattern of biomarker correlations in participants who died was consistent with that of the entire cohort, with positive pairwise correlations found among G-CSF, GM-CSF, IFN-γ, IL-1β, IL-4, IL-5, IL-12, IL-13, IL-17, MCP1MCAF, and TNF-α **(Figure S16)**. Negative pairwise correlations included ApoB100 IC IgA with MCP1MCAF and anti-P1 IgM with IgA. Clusters were consistent across mortality status, with the most distinct separation between clusters found among survivors **(Figure S7)**. After disaggregation, clusters were similar between participants who died with tuberculosis compared to those who died without tuberculosis **(Figure S7)**. Regardless of tuberculosis status, high concentrations of GM-CSF, IFN-g, IL-1β, IL-4, and TNF-α clustered together. PCA identified IL-4, IL-6, IL-8, IL-12, and IL-13 as most associated with participants who died, and G-CSF, IL-6, IL-8, MIP-1β, and MCP1MCAF as most associated with participants who survived **(Table S1; Figure 1)**.

#### Multivariable logistic regression models of mortality

A multivariable biomarker model that included IL-6, IL-8, IL-10, and MIP-1β predicted mortality with an AUC of 0.72 (95% CI 0.64-0.80) **(Table 2 and Figure 4)**. This biomarker model plus anti-P1 IgM and anti-ApoB100 IC IgM predicted mortality with an AUC of 0.74 (95% CI 0.67-0.81). A comprehensive biomarker and clinical multivariable logistic regression model that added HIV serostatus and lactate to the prior model predicted mortality with an AUC of 0.78 (95% CI 0.70-0.84). The addition of IL-8 and lactate to UVA score improved the discrimination of mortality from 0.57 (95% CI 0.49-0.65) to 0.71 (95% CI 0.63-0.78). We replicated these results in the validation cohort **(Table 2)**. The biomarker models with cytokines and antibodies underpredicted mortality risk when the observed mortality rate was high, while the comprehensive model overestimated mortality risk when the observed mortality rate was low **(Figure 4)**. After disaggregation, the mortality models performed best among participants with tuberculosis **(Table S4)**.

### Cross-testing of multivariable models of tuberculosis and mortality

The comprehensive multivariable logistic regression model built to predict tuberculosis (IL-1β, IL-8, total IgM, HIV serostatus, and blood lactate) predicted mortality with an AUC of 0.77 (95% CI 0.69-0.84) compared to an AUC of 0.84 (95% CI 0.77-0.89) for tuberculosis **(Table 2)**. The comprehensive tuberculosis prediction model underpredicted mortality at higher observed mortality **(Figure 4)**. The comprehensive multivariable logistic regression model built to predict mortality (IL-6, IL-8, IL-10, MIP-1β, anti-P1 IgM, anti-ApoB100IC IgM, HIV serostatus, and lactate) predicted tuberculosis with an AUC of 0.81 (95% CI 0.74-0.87) compared to an AUC of 0.78 (0.70-0.84) for mortality. The comprehensive mortality prediction model underpredicted tuberculosis at the lowest and highest rates of observed tuberculosis **(Figure 4)**. Tuberculosis prediction models performed poorly when predicting participants without tuberculosis, with or without other pathogens, and whether or not they died **(Table S4)**.

## DISCUSSION

In PLWH with sepsis in East Africa, tuberculosis was associated with systemic inflammation marked by chemotaxis and macrophage activation, with G-CSF, IL-5, IL-6, IL-8, and IL-13 identified by PCA as biomarkers with the highest loadings. In the PCA, mortality was associated with hyperinflammatory cytokines, IL-4, IL-6, IL-8, IL-12, and IL-13, and dysregulated Th1/Th2 signaling through IL-4, IL-12, and IL-13. Comprehensive biomarker and clinical multivariable models accurately predicted tuberculosis and mortality. IL-8 was the single most predictive biomarker for both tuberculosis and mortality, while the anti-OSE antibody anti-ApoB100 IC IgM had a consistent inverse association with mortality regardless of tuberculosis status. Immune responses were independent of CD4+ T-cell and lactate concentrations. The results from the discovery cohort were replicated in the validation cohort.

In both discovery and validation cohorts, tuberculosis was associated with increased inflammation regardless of mortality outcomes as measured by individual biomarkers and the ratios of pro-inflammatory cytokines to IL-10. Predictive models of tuberculosis that included IL-1β and IL-8 performed poorly among participants without tuberculosis who were infected with a pathogen other than tuberculosis, whether or not they died. This finding suggests that tuberculosis leads to a unique immune subphenotype. In separate studies of adult sepsis in Uganda, HIV-associated disseminated tuberculosis was similarly associated with a pro-inflammatory immune subphenotype that included IFN-γ, IL-6, IL-8, MIF, MIP-1β, and TNF-α (21–23).

Collectively, our findings suggest that the immune perturbations underpinning tuberculosis and death in sepsis are overlapping but not identical, indicating shared immune pathways. The comprehensive tuberculosis model predicted tuberculosis with an AUC of 0.84 and mortality with an AUC of 0.77, whereas the comprehensive mortality model predicted mortality with an AUC of 0.78 and tuberculosis with an AUC of 0.81. The smaller difference between AUCs for the mortality model likely reflects a broader signature of immune dysregulation that encompassed both tuberculosis and non-tuberculosis inflammatory responses. In contrast, the tuberculosis model identified immune features characteristic of tuberculosis that were less generalizable to mortality.

We found that a pro-inflammatory subphenotype including G-CSF, IL-6, IL-8, IL-10, MCP1MCAF, and MIP-1β was associated with increased mortality. This finding is consistent with other studies that showed that a hyperinflammatory response is detrimental in severely-ill patients with HIV (24, 25). In a prior study of sepsis in Uganda, patients had a transcriptomic sepsis response signature of innate immune activation and T-/NK-cell exhaustion that was associated with higher mortality (25). In our study, the pro-inflammatory subphenotype associated with mortality was consistent across both the discovery and validation cohorts and may reflect a host response aimed at controlling and eliminating pathogens through the recruitment of innate immune cells including neutrophils, macrophages, and natural killer cells (26).

Sepsis is associated with increased oxidative stress and increased formation of OSEs, which promote immune activation and inflammation (13). Anti-OSE antibodies bind to OSEs and anti-OSE IgM can limit inflammation in atherosclerosis (27, 28). We observed that anti-OSE antibodies, anti-P1 IgM, anti-P1 IgG, and anti-ApoB100 IC IgM, were associated with decreased 30-day mortality. Since these antibodies are present across species without prior antigenic stimulation, we hypothesize that they represent an evolutionary conserved immunity that limits inflammation and tissue damage in sepsis. Rather than relying on a pro-inflammatory host response to eliminate pathogens, anti-OSE antibody expression may promote disease tolerance, a strategy that preserves tissue integrity and physiological function without directly targeting the pathogen (26).

In general, biomarkers should augment disease screening, diagnosis, risk stratification, or monitoring; or serve as a surrogate endpoint (29). In our study, a comprehensive model combining biomarkers and clinical variables improved prediction of tuberculosis and 30-day mortality, and a combination of cytokines and anti-OSE antibodies predicted mortality with good performance. While we acknowledge that a multi-biomarker model is likely not feasible for rapid identification of high-risk patients, we also found that adding only IL-8 together with lactate significantly improved UVA score performance. These findings suggest that IL-8 might be of clinical utility in diagnosing patients with HIV-related tuberculosis sepsis and a high-risk, hyperinflammatory subphenotype.

This study had several limitations. First, the discovery and validation cohorts predominantly comprise PLWH enrolled across a decade at a limited number of hospitals in East Africa, which may restrict the generalizability of the findings to populations with different comorbidities, healthcare resources, or epidemiologic contexts across Africa. Second, biomarker measurements were obtained at a single time point, precluding assessment of dynamic immune changes over the course of illness (30). Third, although the study identified associations between biomarkers, tuberculosis, and mortality, causality cannot be inferred. Fourth, potential confounding from unmeasured factors such as coinfections, treatment heterogeneity, undernutrition, or differences in pathogen burden could have influenced the observed relationships. Finally, while the sample size was sufficient to detect associations and was externally validated in a similar cohort, external validation in larger, geographically diverse cohorts is needed to confirm the reproducibility and clinical utility of these immune subphenotypes.

To conclude, among adults with HIV-related sepsis in East Africa, tuberculosis was associated with a distinct pro-inflammatory immune subphenotype marked by chemotaxis, and neutrophil and macrophage activation, whereas mortality reflected a broader hyperinflammatory state and dysregulated Th1/Th2 signaling. Our findings align with multi-omics analyses, which identified hyperactivated T-cell responses with monocyte and neutrophil-driven IFN and IL-1-mediated inflammation in disseminated tuberculosis (31). Elevated G-CSF, IL-6, IL-8, IL-10, MCP1MCAF, and MIP-1β were associated with increased mortality, while anti-OSE antibodies were associated with survival, which delineates complementary pro-inflammatory patterns of host response and disease tolerance. IL-8 was the single biomarker most associated with both tuberculosis and mortality. Together, these findings highlight shared but distinct immune pathways of tuberculosis and death in HIV-related sepsis and support the use of immune subphenotypes to improve risk-stratification and guide targeted interventions.

## Data Availability

All data produced in the present study are available upon reasonable request to the authors

## ACKNOWLEDGEMENTS

The PRISM-U2 trial was supported by an Investigator-Initiated Award provided by Pfizer, Inc. The ATLAS trial was supported by the National Institutes of Health (U01 AI150508 to SKH and CCM; D43 TW012247 to SKH; R21 AI172637 to TAT; K24 AI187675 to SKH). The anti-OSE assays were supported by the National Institutes of health (R01 HL179312 to JMS), the Wetmore Foundation (25-0018-P0001 to EO), and Tulane University (Tulane Physician Scientist Pipeline Program Scholar Award to EO). The funders had no role in the design and conduct of the study; the collection, management, analysis and interpretation of the data; or the preparation, review or approval of the manuscript.

---

**Supplementary Table 1.**
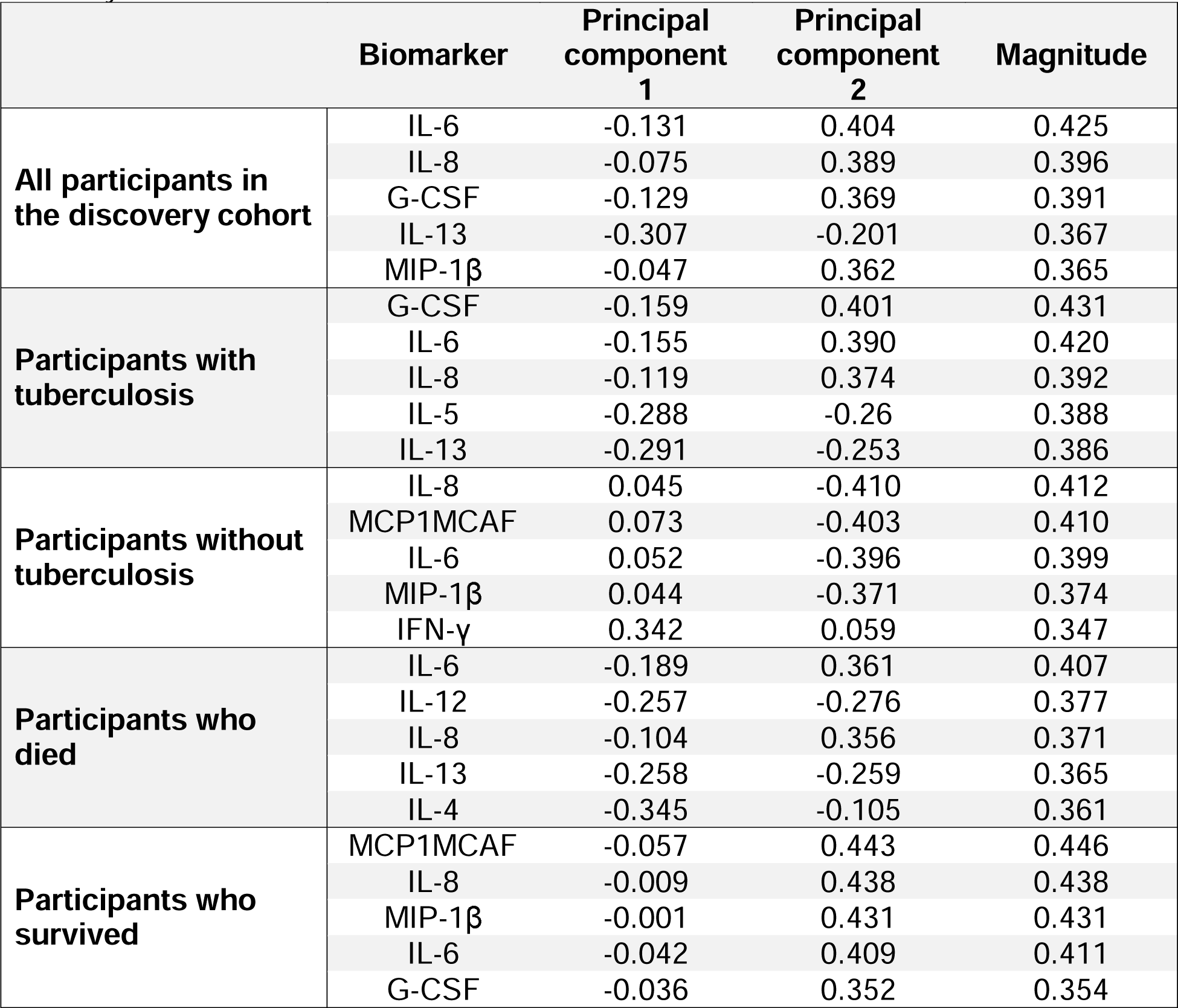
Principal component analysis loadings for the top five biomarkers among adults with sepsis in Uganda in the discovery cohort, stratified by tuberculosis and mortality status.

**Supplementary Table 2.** Odds ratios and 95% confidence intervals for associations between biomarkers and tuberculosis among adults with sepsis in the discovery cohort.

| Predictor | Odds ratio | 95% Confidence interval | P-value | Adjusted P-value |
| --- | --- | --- | --- | --- |
| IL-1β | 2.30 | 1.64-3.24 | <0.00001 | 0.00004 |
| IgM | 1.81 | 1.05-3.11 | 0.03 | 0.21 |
| IL-8 | 1.60 | 1.23-2.09 | 0.0005 | 0.007 |
| IL-7 | 1.59 | 0.93-2.72 | 0.09 | 0.40 |
| MCP1MCAF | 1.37 | 1.11-1.70 | 0.004 | 0.032 |
| IL-13 | 1.34 | 0.84-2.15 | 0.22 | 0.52 |
| IFN-γ | 1.28 | 0.91-1.80 | 0.15 | 0.52 |
| Anti-ApoB100 IC IgM | 1.25 | 0.80-1.96 | 0.32 | 0.59 |
| TNF-α | 1.25 | 0.85-1.83 | 0.25 | 0.52 |
| IgA | 1.25 | 0.86-1.81 | 0.24 | 0.52 |
| MIP-1β | 1.22 | 0.86-1.74 | 0.26 | 0.52 |
| IL-6 | 1.22 | 0.99-1.50 | 0.06 | 0.30 |
| G-CSF | 1.12 | 0.92-1.36 | 0.26 | 0.52 |
| GM-CSF | 1.08 | 0.75-1.56 | 0.66 | 0.87 |
| IL-4 | 1.08 | 0.67-1.75 | 0.74 | 0.87 |
| Anti-ApoB100 IgA | 1.08 | 0.64-1.81 | 0.77 | 0.87 |
| Anti-P1 IgA | 1.04 | 0.70-1.53 | 0.85 | 0.91 |
| Anti-ApoB100 IgG | 1.03 | 0.60-1.76 | 0.91 | 0.91 |
| IL-2 | 1.02 | 0.75-1.40 | 0.89 | 0.91 |
| IL-10 | 0.95 | 0.73-1.23 | 0.69 | 0.87 |
| IL-12 | 0.91 | 0.48-1.70 | 0.76 | 0.87 |
| IL-17 | 0.88 | 0.48-1.62 | 0.68 | 0.87 |
| Anti-P1 IgM | 0.87 | 0.58-1.33 | 0.53 | 0.86 |
| IgG | 0.86 | 0.50-1.47 | 0.58 | 0.87 |
| IL-5 | 0.79 | 0.45-1.39 | 0.42 | 0.72 |
| Anti-P1 IgG | 0.74 | 0.46-1.17 | 0.19 | 0.52 |

**Supplementary Table 3.** Odds ratios and 95% confidence intervals for associations between biomarkers and mortality among adults with sepsis in the discovery cohort.

| Predictor | Odds ratio | 95% Confidence interval | P-value | Adjusted P-value |
| --- | --- | --- | --- | --- |
| IL-8 | 1.92 | 1.44-2.56 | 0.00001 | 0.0002 |
| MIP-1β | 1.66 | 1.18-2.35 | 0.004 | 0.02 |
| MCP1MCAF | 1.54 | 1.24-1.92 | 0.0001 | 0.001 |
| IL-6 | 1.48 | 1.20-1.82 | 0.0002 | 0.002 |
| G-CSF | 1.34 | 1.10-1.64 | 0.004 | 0.02 |
| IL-1β | 1.34 | 1.01-1.76 | 0.04 | 0.10 |
| IL-4 | 1.31 | 0.82-2.11 | 0.25 | 0.51 |
| IFN-γ | 1.30 | 0.93-1.82 | 0.13 | 0.28 |
| IL-10 | 1.29 | 1.02-1.63 | 0.04 | 0.10 |
| IL-13 | 1.25 | 0.80-1.96 | 0.33 | 0.60 |
| IL-17 | 1.24 | 0.69-2.23 | 0.46 | 0.65 |
| IgM | 1.19 | 0.75-1.91 | 0.45 | 0.65 |
| IL-7 | 1.16 | 0.69-1.96 | 0.57 | 0.70 |
| TNF-γ | 1.12 | 0.77-1.61 | 0.55 | 0.70 |
| IgG | 1.09 | 0.65-1.82 | 0.75 | 0.81 |
| GM-CSF | 1.07 | 0.75-1.52 | 0.71 | 0.81 |
| IL-12 | 1.00 | 0.57-1.75 | 1.00 | 1.00 |
| IgA | 0.97 | 0.69-1.37 | 0.87 | 0.90 |
| IL-2 | 0.92 | 0.68-1.26 | 0.62 | 0.73 |
| Anti-P1 IgA | 0.85 | 0.59-1.24 | 0.41 | 0.65 |
| Anti-ApoB100 IC IgA | 0.84 | 0.52-1.35 | 0.47 | 0.65 |
| IL-5 | 0.79 | 0.47-1.33 | 0.38 | 0.65 |
| Anti-ApoB100 IC IgG | 0.65 | 0.38-1.11 | 0.11 | 0.27 |
| Anti-P1 IgM | 0.61 | 0.40-0.94 | 0.024 | 0.079 |
| Anti-P1 IgG | 0.57 | 0.36-0.89 | 0.01 | 0.05 |
| Anti-ApoB100 IC IgM | 0.56 | 0.36-0.86 | 0.008 | 0.04 |

**Supplementary Table 4.** Multinomial multivariate logistic regression models for the prediction of tuberculosis or other pathogens among adults with sepsis in the discovery cohort.

| Discovery cohort<br>AUC (95% confidence interval) |  |  |  |  |  |  |
| --- | --- | --- | --- | --- | --- | --- |
|  | Participants who died |  |  | Participants who survived |  |  |
|  | TB+ | TB- OP+ | TB- OP- | TB+ | TB- OP+ | TB- OP- |
| Age + Sex + IL-1β + IL-8 | 0.88<br>(0.82-0.93) | 0.62<br>(0.46-0.77) | 0.79<br>(0.65-0.88) | 0.76<br>(0.69-0.83) | 0.64<br>(0.52-0.74) | 0.68<br>(0.58-0.76) |
| Age + Sex + IL-1β + IL-8 + IgM | 0.90<br>(0.84-0.95) | 0.62<br>(0.46-0.78) | 0.80<br>(0.68-0.91) | 0.76<br>(0.69-0.83) | 0.69<br>(0.59-0.78) | 0.71<br>(0.62-0.79) |
| Age + Sex + IL-1β + IL-8 + IgM + Lactate + HIV Status | 0.90<br>(0.84-0.95) | 0.72<br>(0.55-0.85) | 0.83<br>(0.72-0.92) | 0.79<br>(0.71-0.86) | 0.73<br>(0.63-0.81) | 0.74<br>(0.65-0.81) |

TB+, confirmed tuberculosis; TB-, no tuberculosis; OP+, other pathogen detected; OP-, no other pathogen detected.

## SUPPLEMENTARY FIGURE LEGENDS

**Supplementary figure 1.**
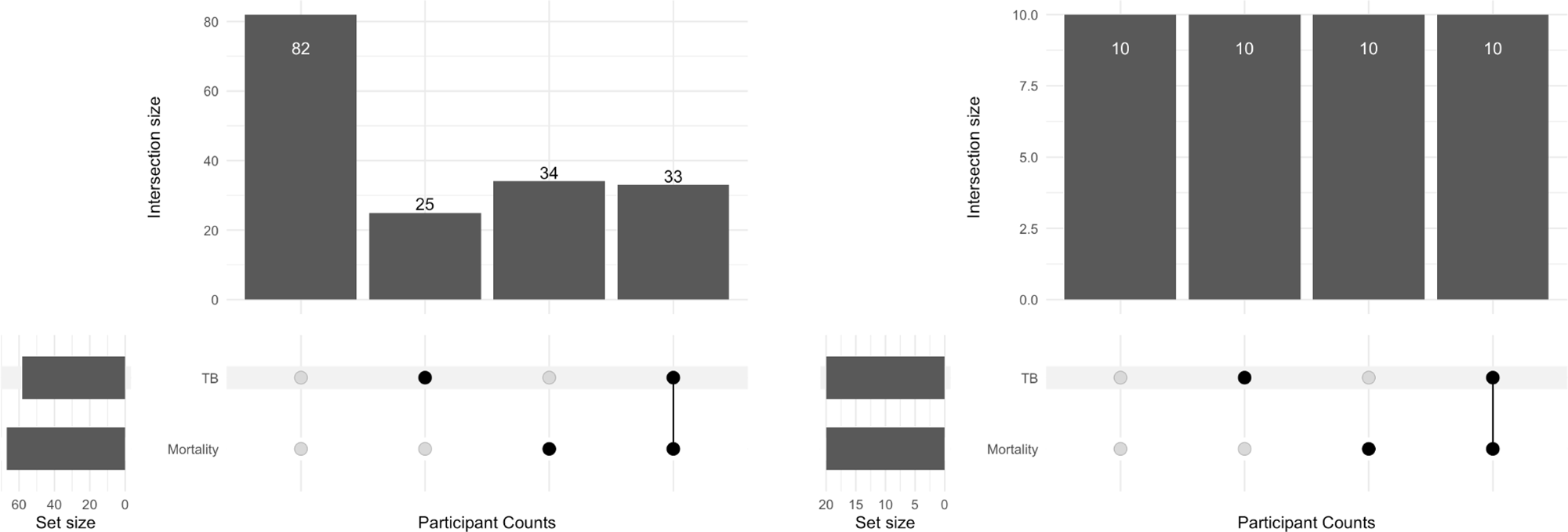
Upset plots of participant tuberculosis and mortality status in the discovery and validation cohorts.

**Supplementary figure 2.**
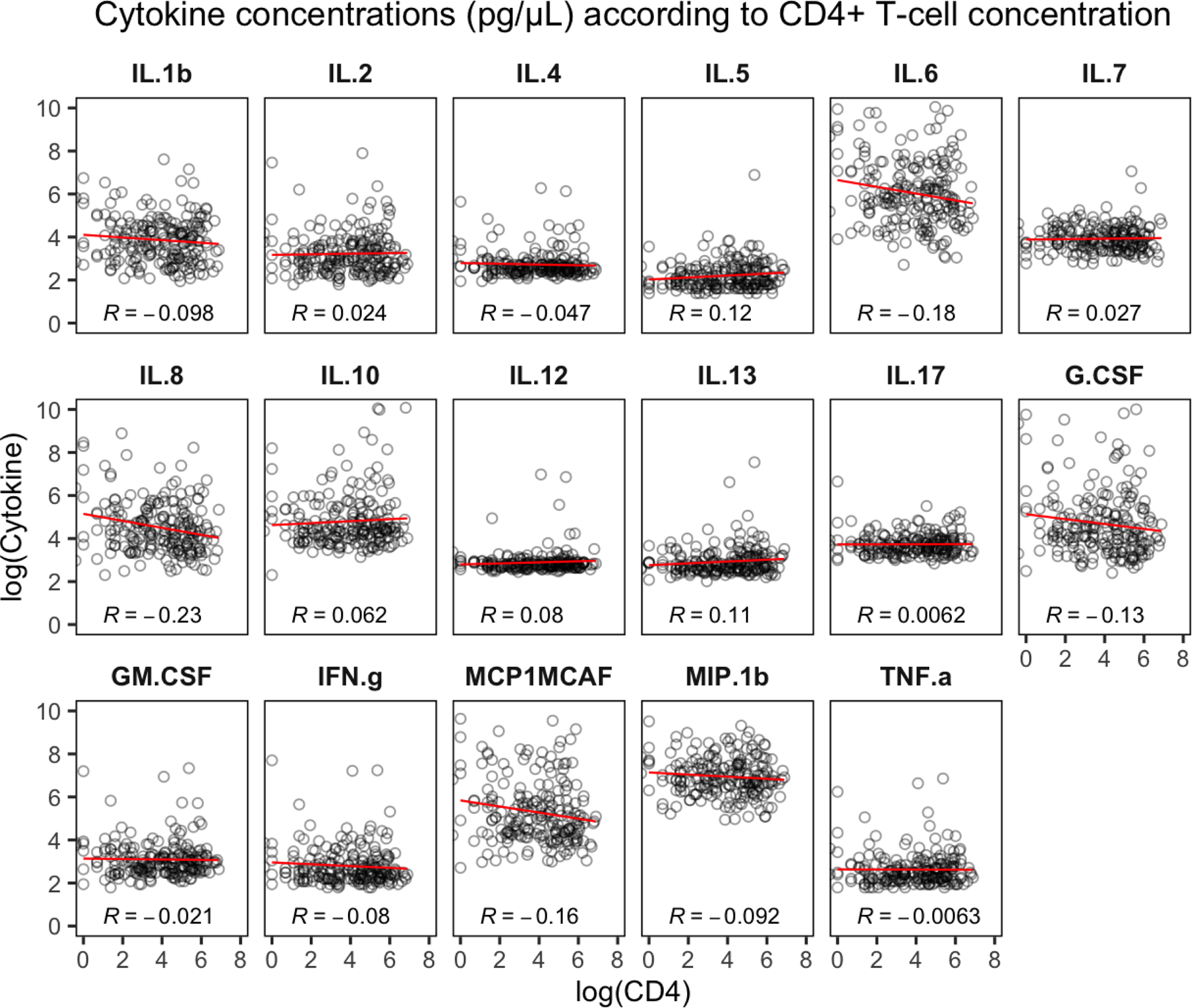
Cytokine concentrations according to log CD4+ T-cell concentration among adults with sepsis in Uganda included in the discovery cohort.

**Supplementary figure 3.**
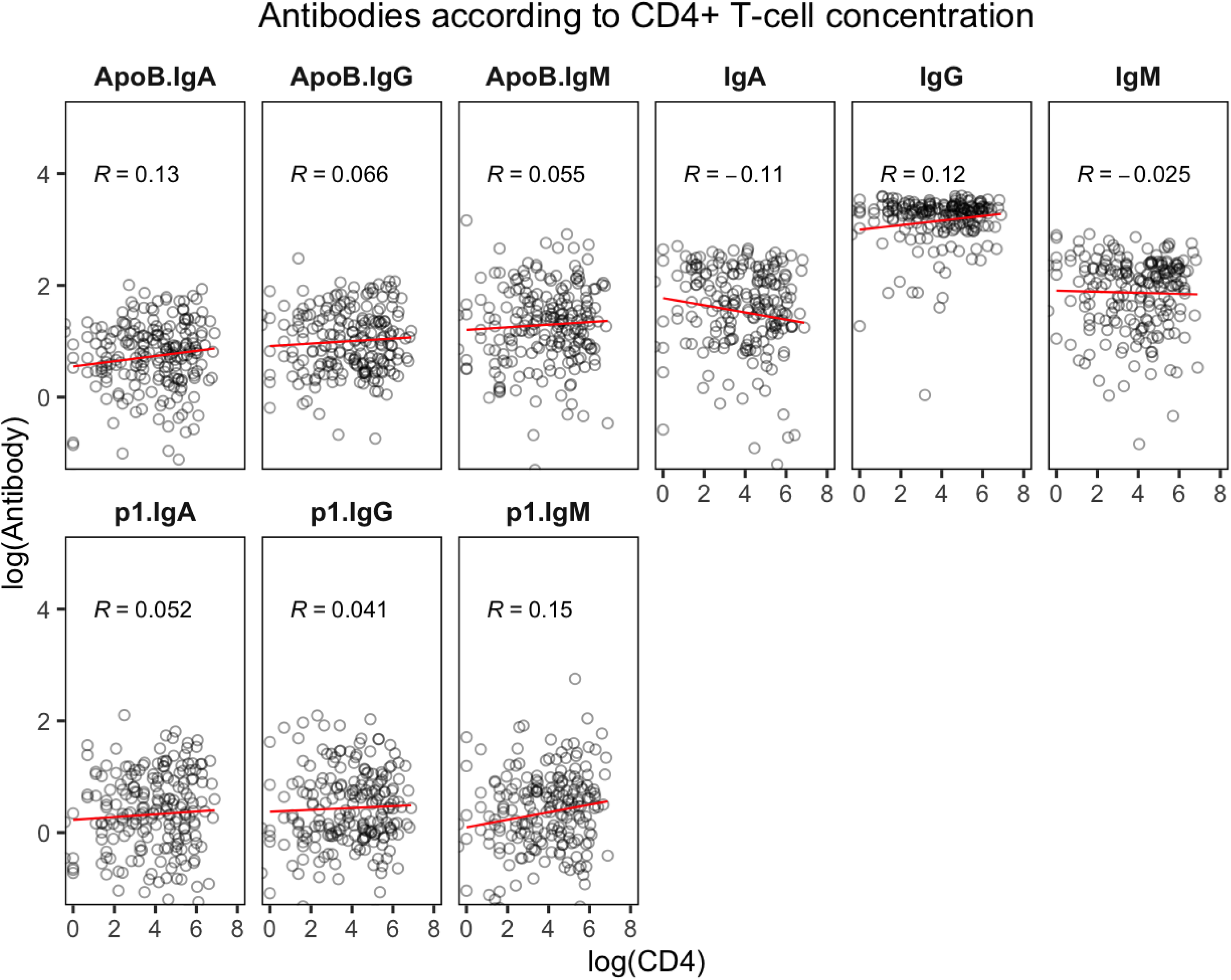
Antibody concentrations according to log CD4+ T-cell concentration among adults with sepsis in Uganda included in the discovery cohort.

**Supplementary figure 4.**
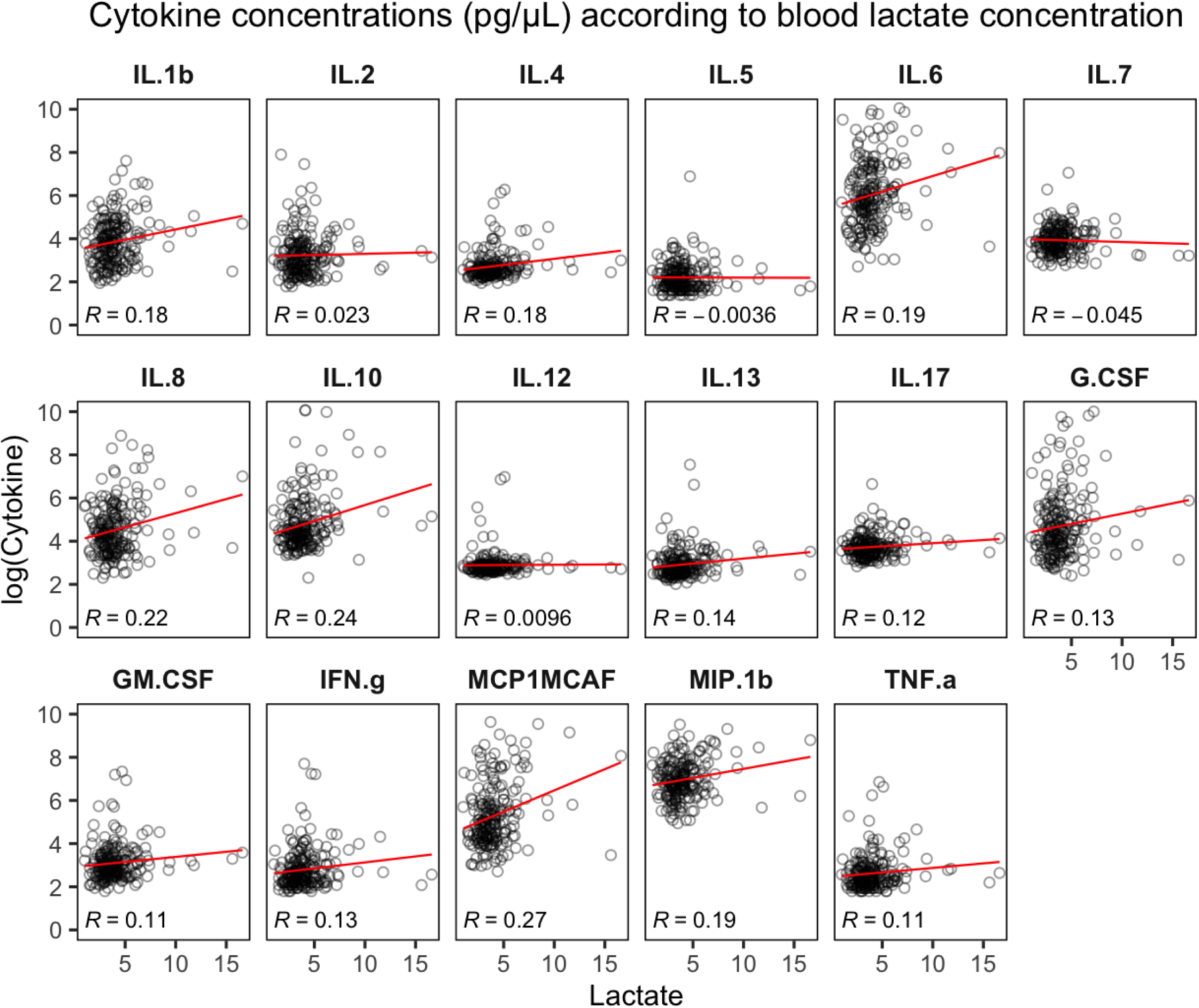
Cytokine concentrations according to whole blood lactate concentration among adults with sepsis in Uganda included in the discovery cohort.

**Supplementary figure 5.**
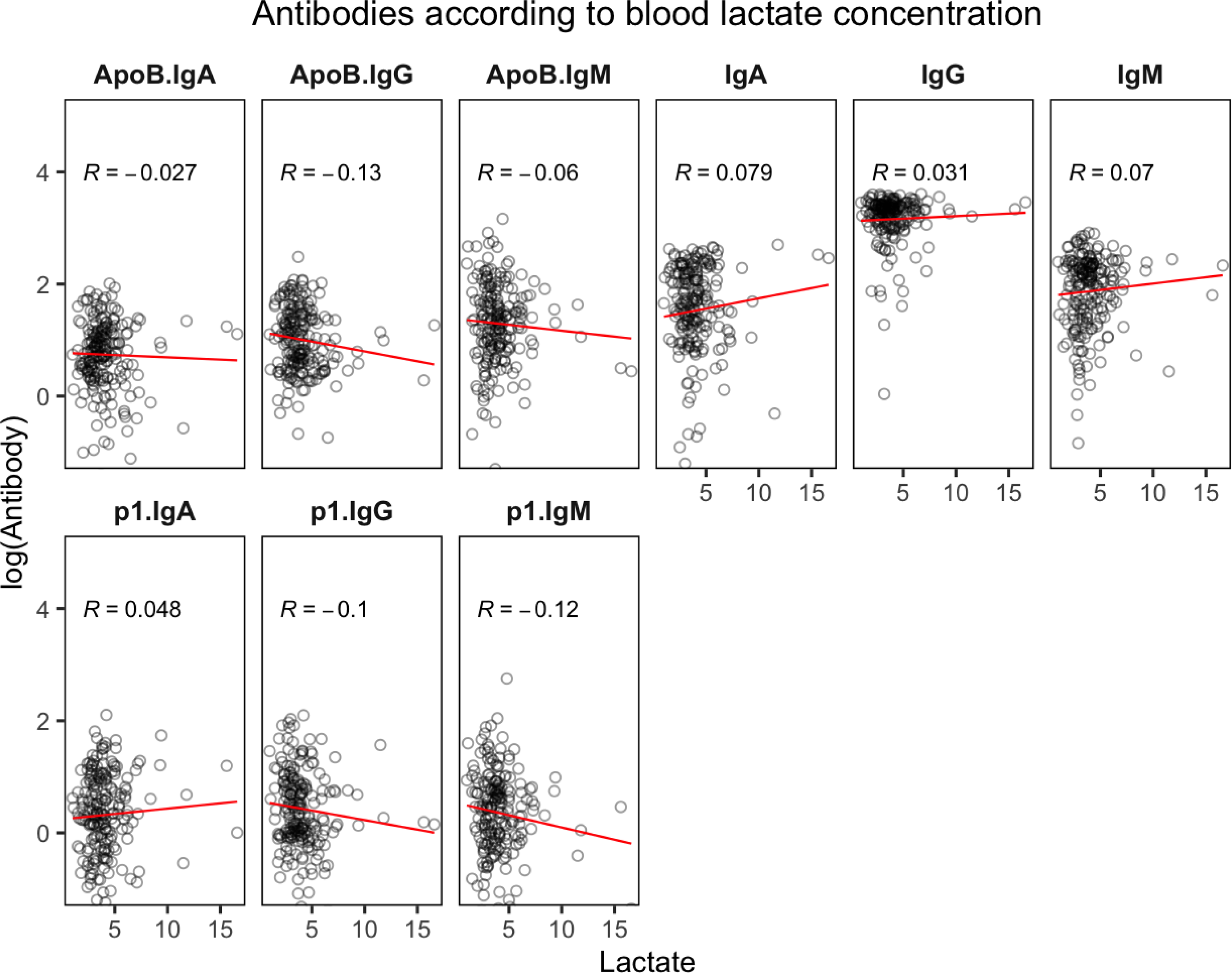
Antibody concentrations according to whole blood lactate concentration (mmol/L) among adults with sepsis in Uganda included in the discovery cohort.

**Supplementary figure 6.**
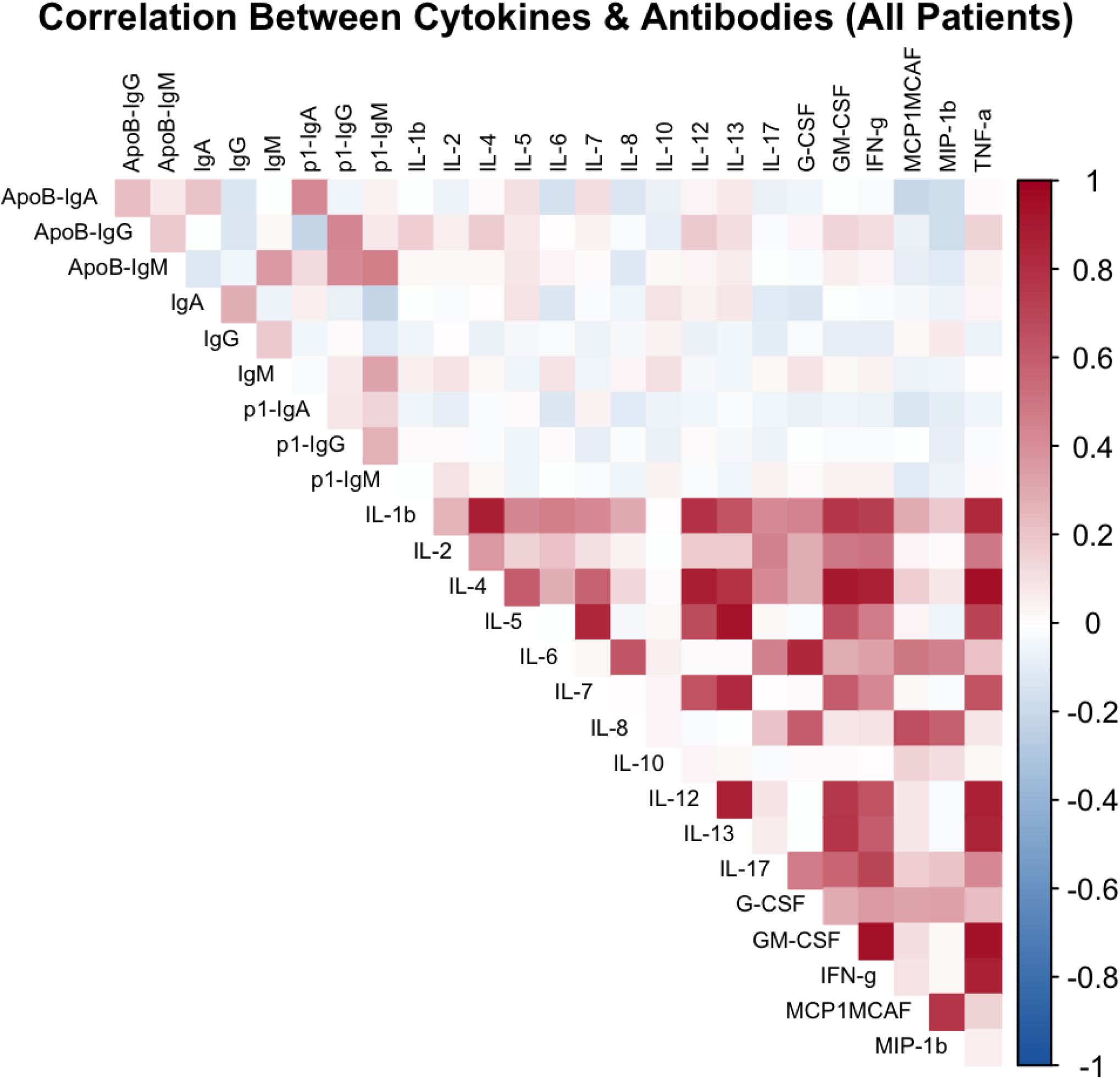
Correlation plot of cytokine and antibody concentrations among adults with sepsis in Uganda included in the discovery cohort. The heatmap values represent Pearson correlation coefficients between pairs of biomarkers.

**Supplementary Figure 7.**
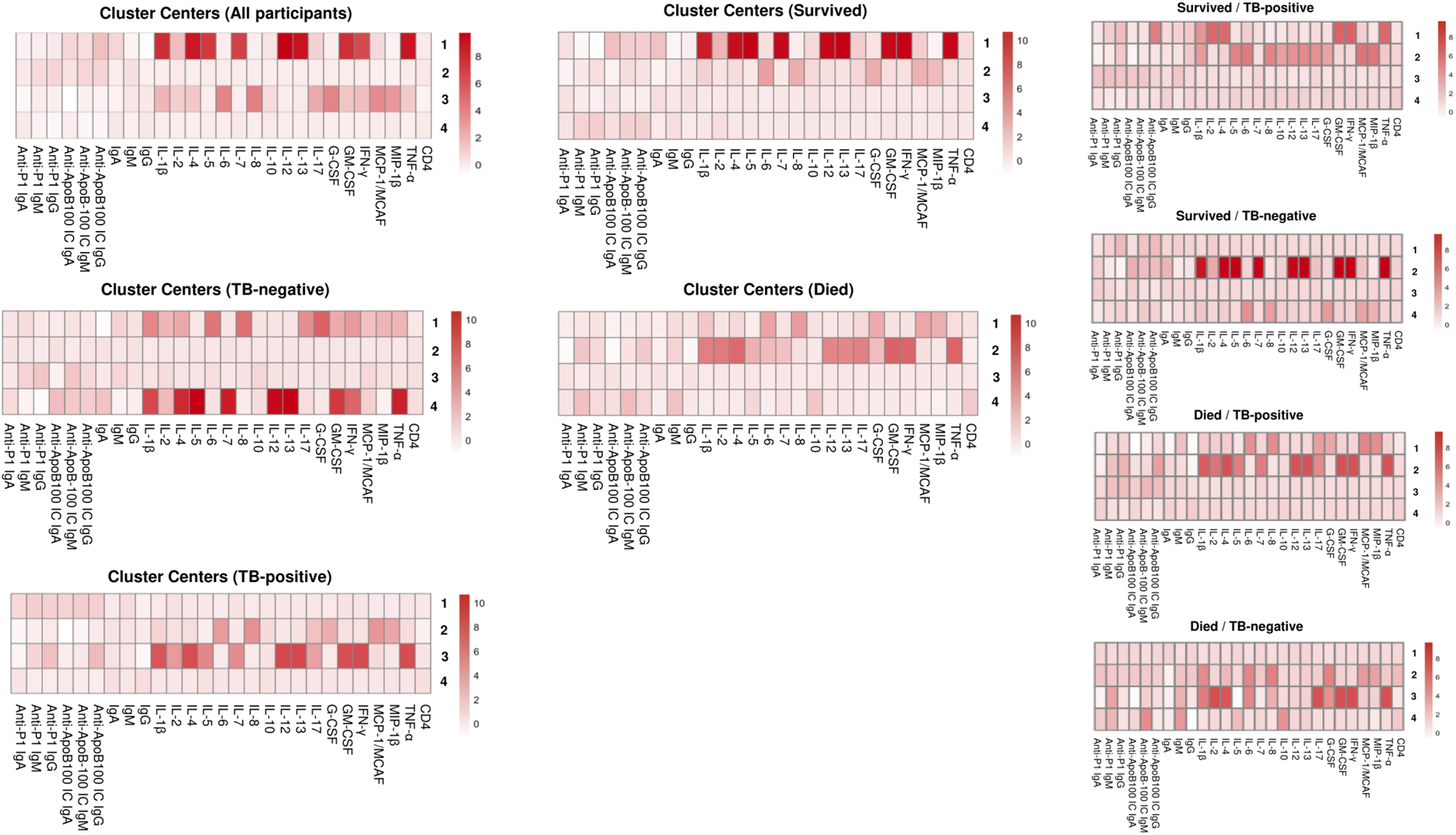
K-means clustering of biomarker profiles among adults with sepsis in Uganda included in the discovery cohort stratified by tuberculosis and mortality status. Clusters represent a distinct group of participants with similar biomarker patterns based on Euclidean distance. The heatmaps display the standardized feature values of the cluster centroids (i.e., average values within each cluster), with darker red shading indicating higher standardized values.

**Supplementary figure 8.**
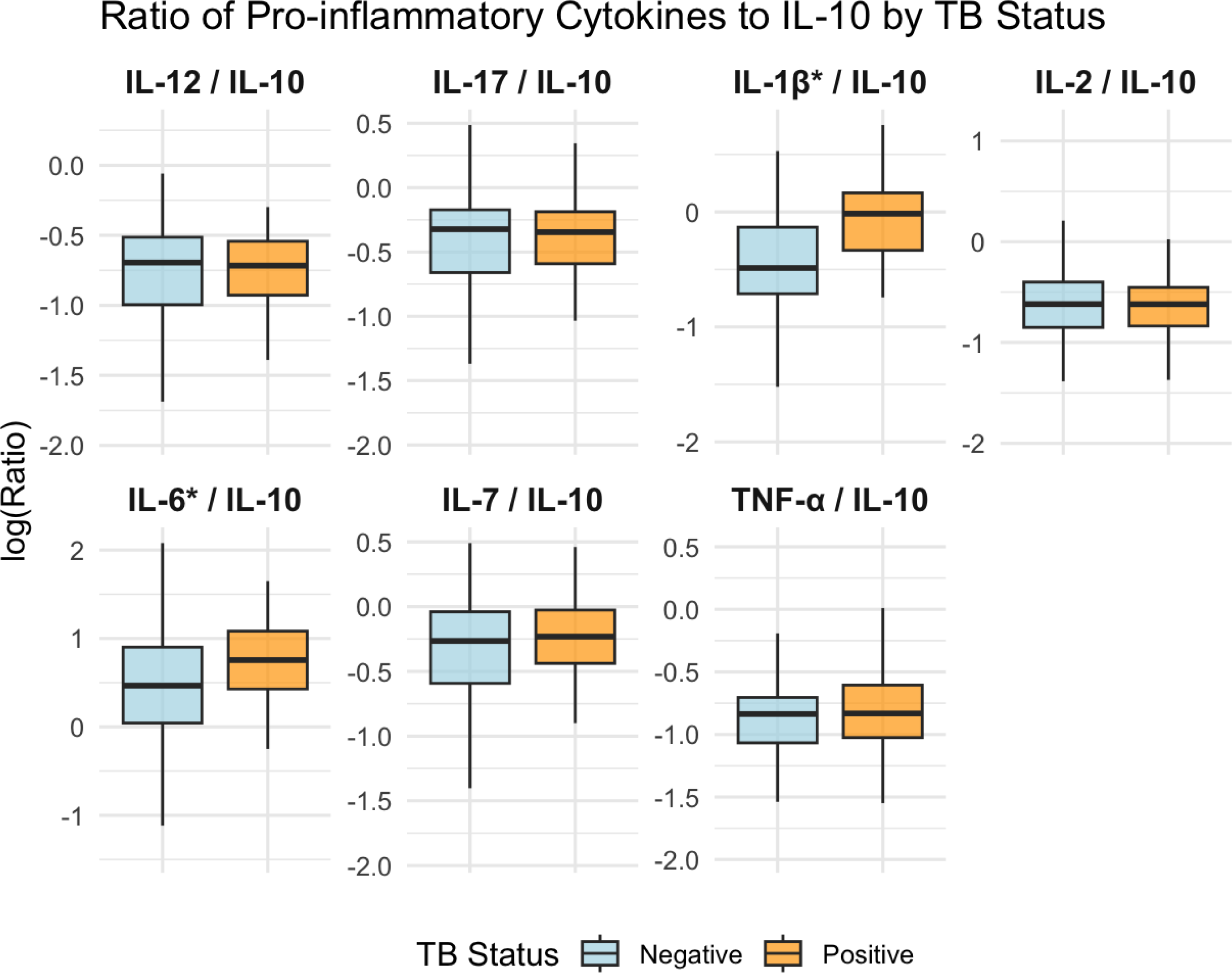
Box-and-whisker plots showing the median, interquartile range, and overall distribution of log-transformed ratios of pro-inflammatory cytokines to interleukin-10 among adults with sepsis in Uganda included in the discovery cohort, stratified by tuberculosis status.

**Supplementary figure 9.**
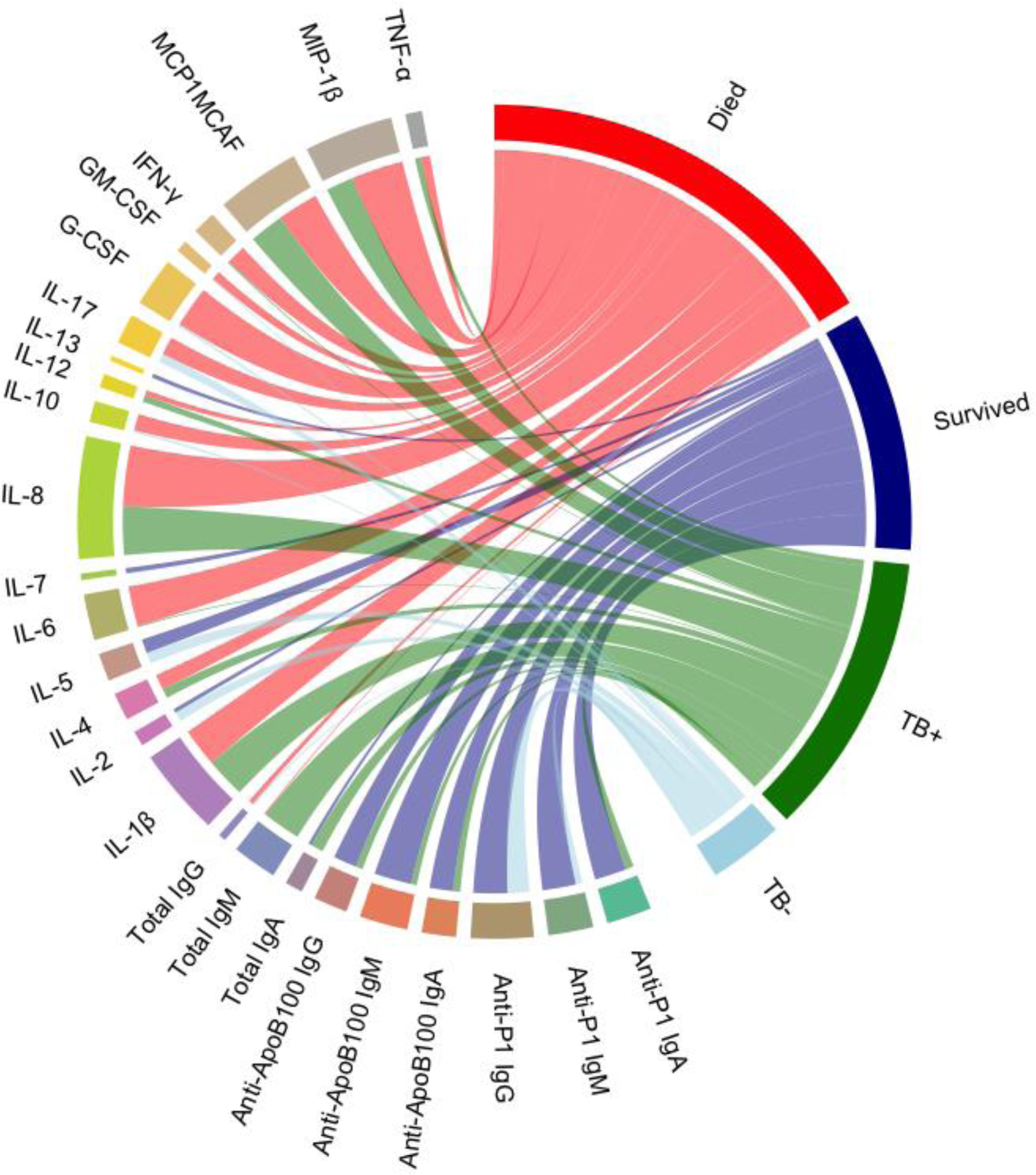
Chord plot showing positive point-biserial correlations between biomarkers, tuberculosis, and 30-day mortality among adults with sepsis in Uganda in the discovery cohort. Chord widths represent the strength of positive correlations, indicating biomarkers elevated in each outcome group. Node widths represent the sums of positive correlations. Larger outcome nodes indicate that biomarkers more strongly discriminate that outcome compared with others.

**Supplementary figure 10.**
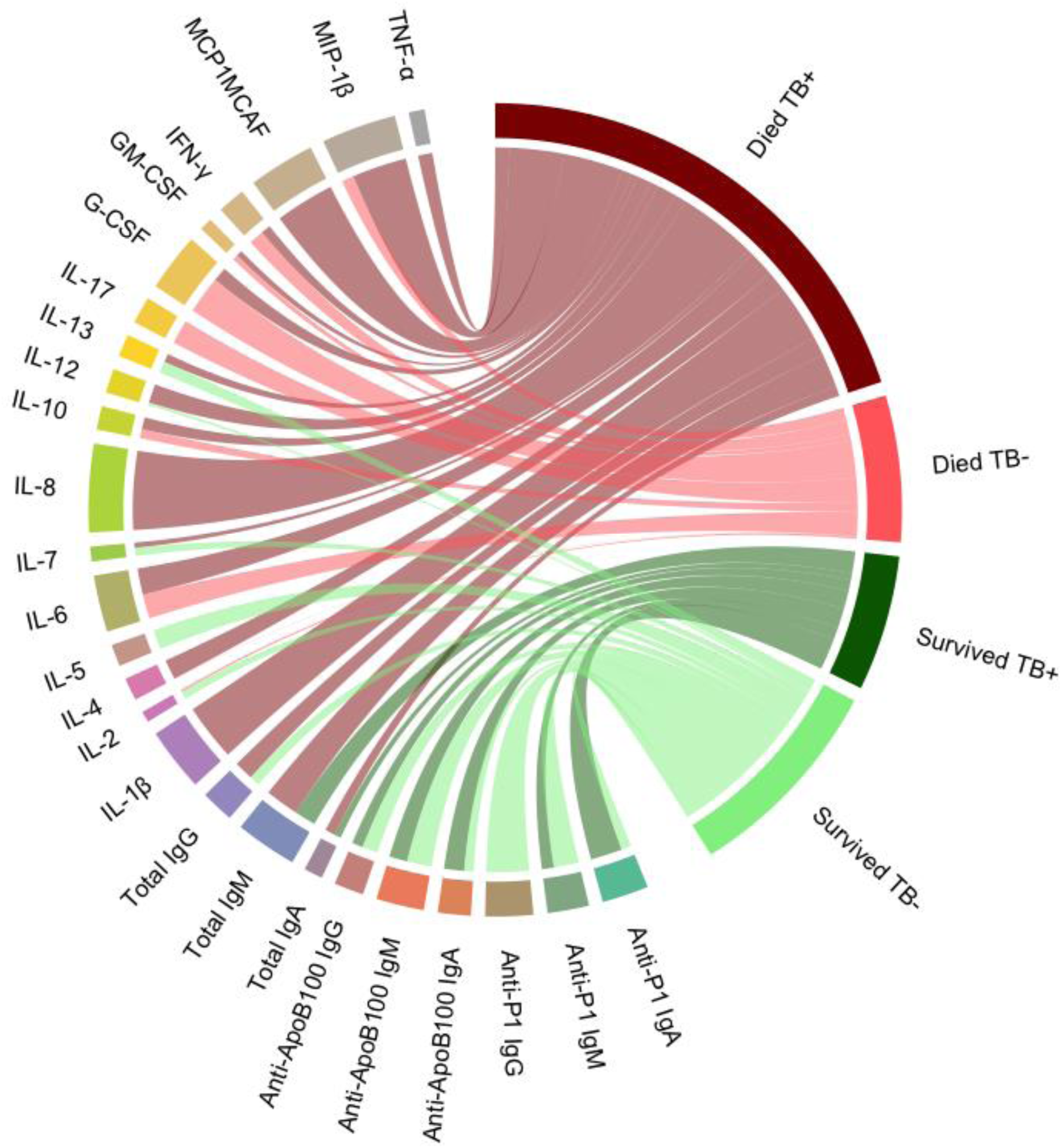
Chord plot showing positive point-biserial correlations between biomarkers and clinical subgroups among adults with sepsis in Uganda included in the discovery cohort. Chord widths represent the strength of positive correlations, indicating biomarkers elevated in each outcome group. Node widths represent the sums of positive correlations. Larger outcome nodes indicate that biomarkers more strongly discriminate that outcome compared with others.

**Supplementary figure 11.**
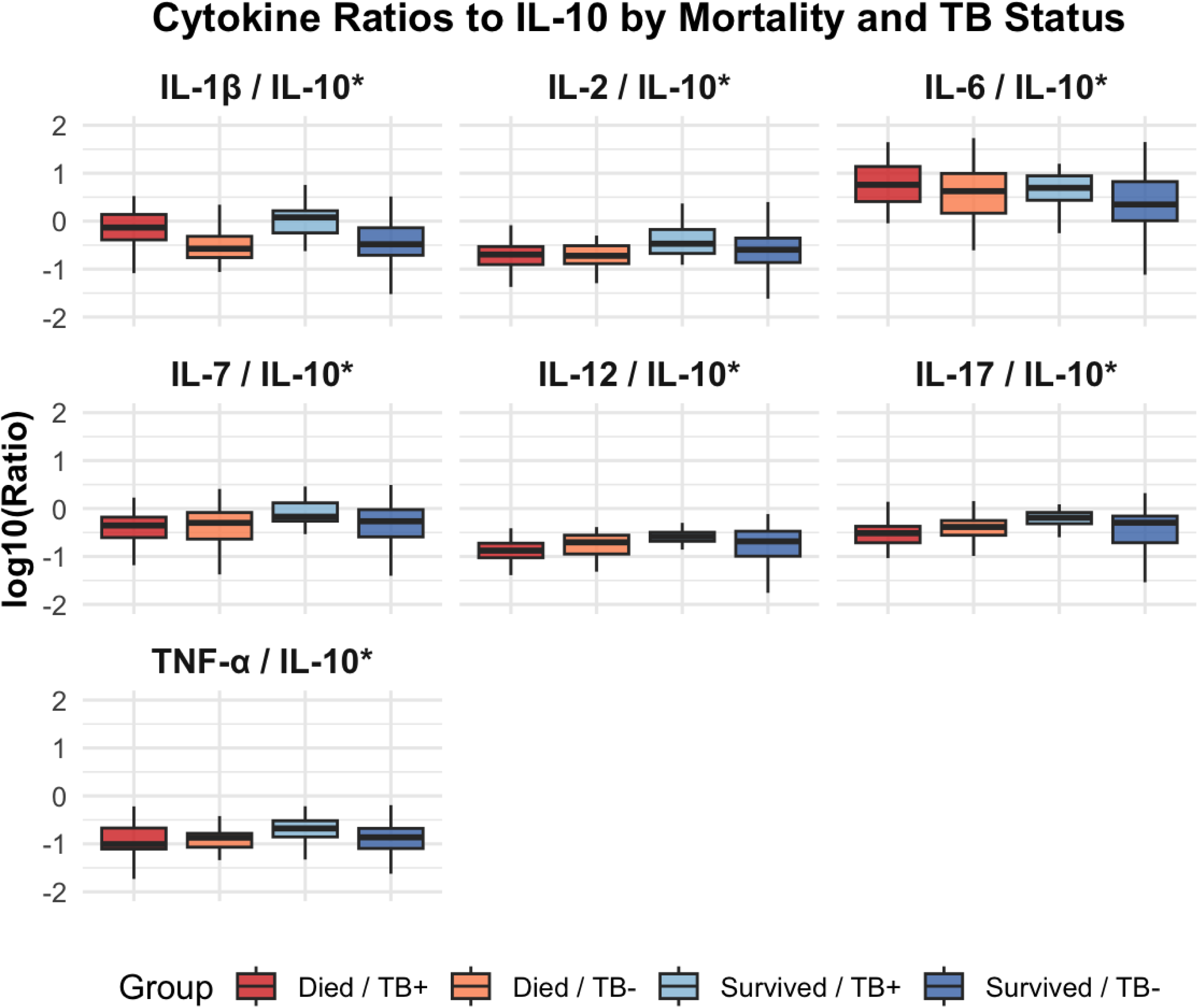
Box-and-whisker plots showing the median, interquartile range, and overall distribution of log-transformed ratios of pro-inflammatory cytokines to interleukin-10 among adults with sepsis in Uganda included in the discovery cohort, stratified by tuberculosis and mortality status.

**Supplementary figure 12.**
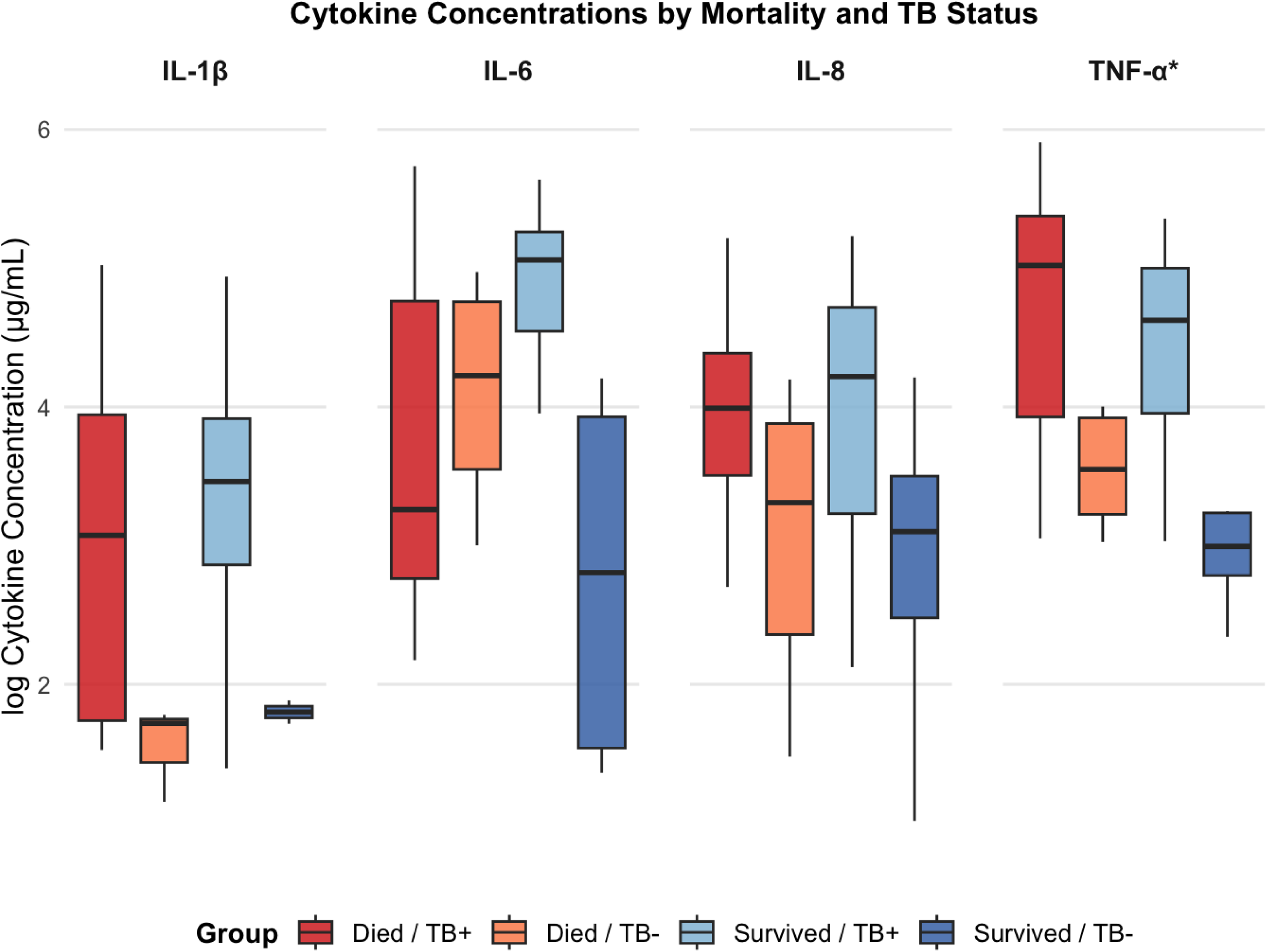
Box-and-whisker plots showing the median, interquartile range, and overall distribution of log-transformed cytokine concentrations among adults with sepsis in Uganda included in the validation cohort, stratified by tuberculosis and mortality status.

**Supplementary figure 13.**
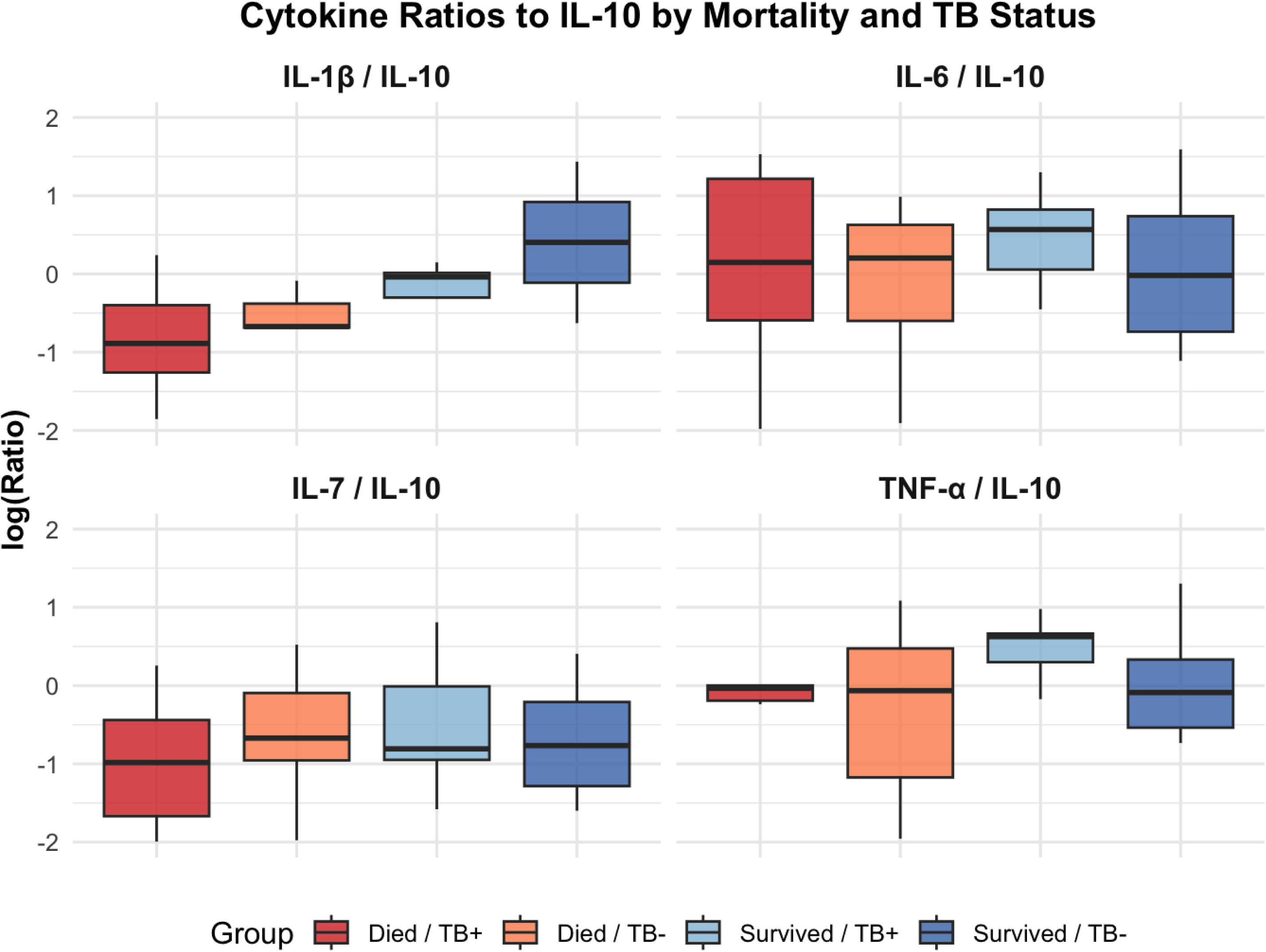
Box-and-whisker plots showing the median, interquartile range, and overall distribution of log-transformed ratios of pro-inflammatory cytokines to interleukin-10 among adults with sepsis in Uganda included in the validation cohort, stratified by tuberculosis and mortality status.

**Supplementary figure 14.**
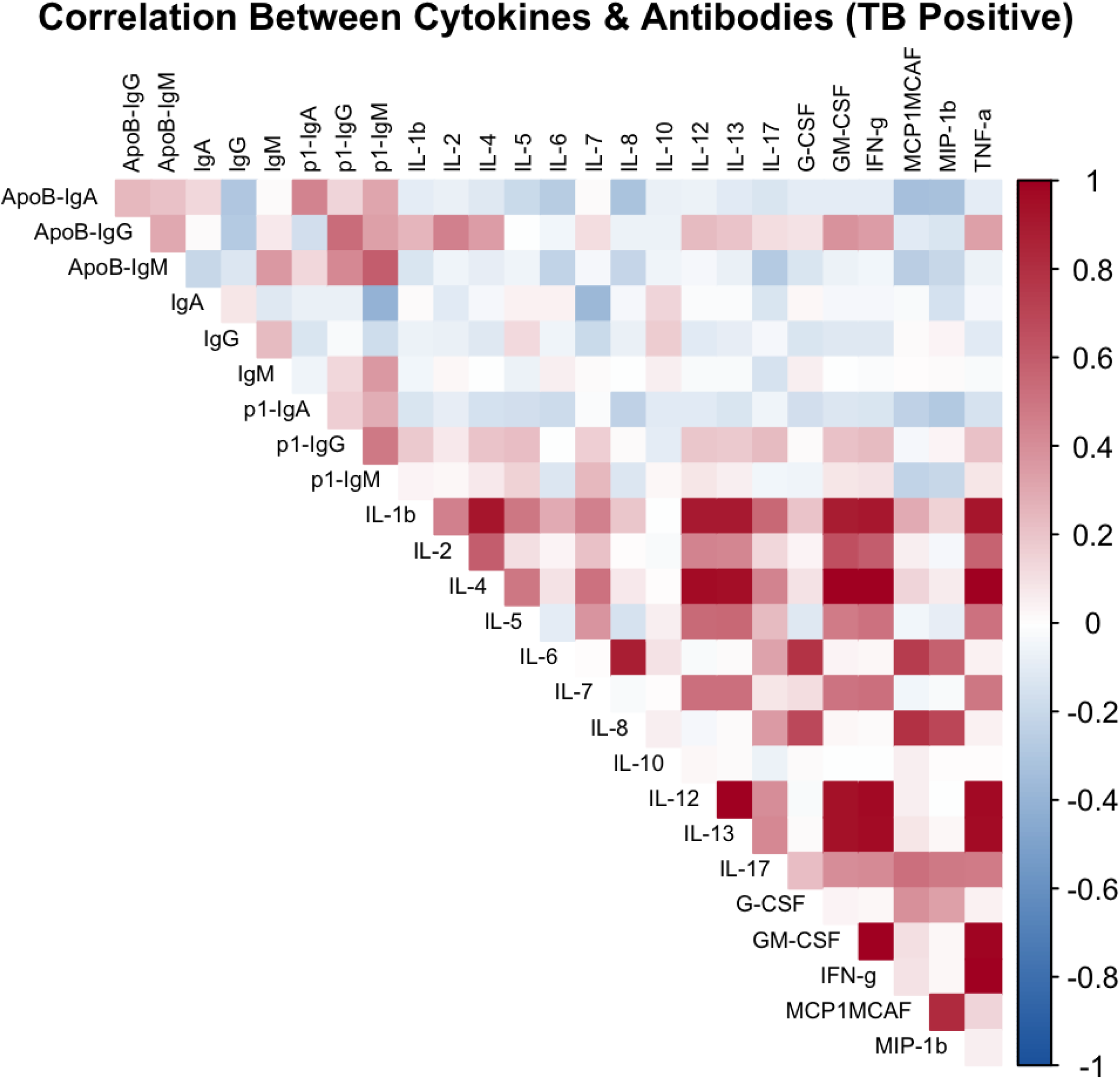
Correlation plot of cytokine and antibody concentrations among adults with sepsis in Uganda included in the discovery cohort with tuberculosis. The heatmap values represent Pearson correlation coefficients between pairs of biomarkers.

**Supplementary figure 15.**
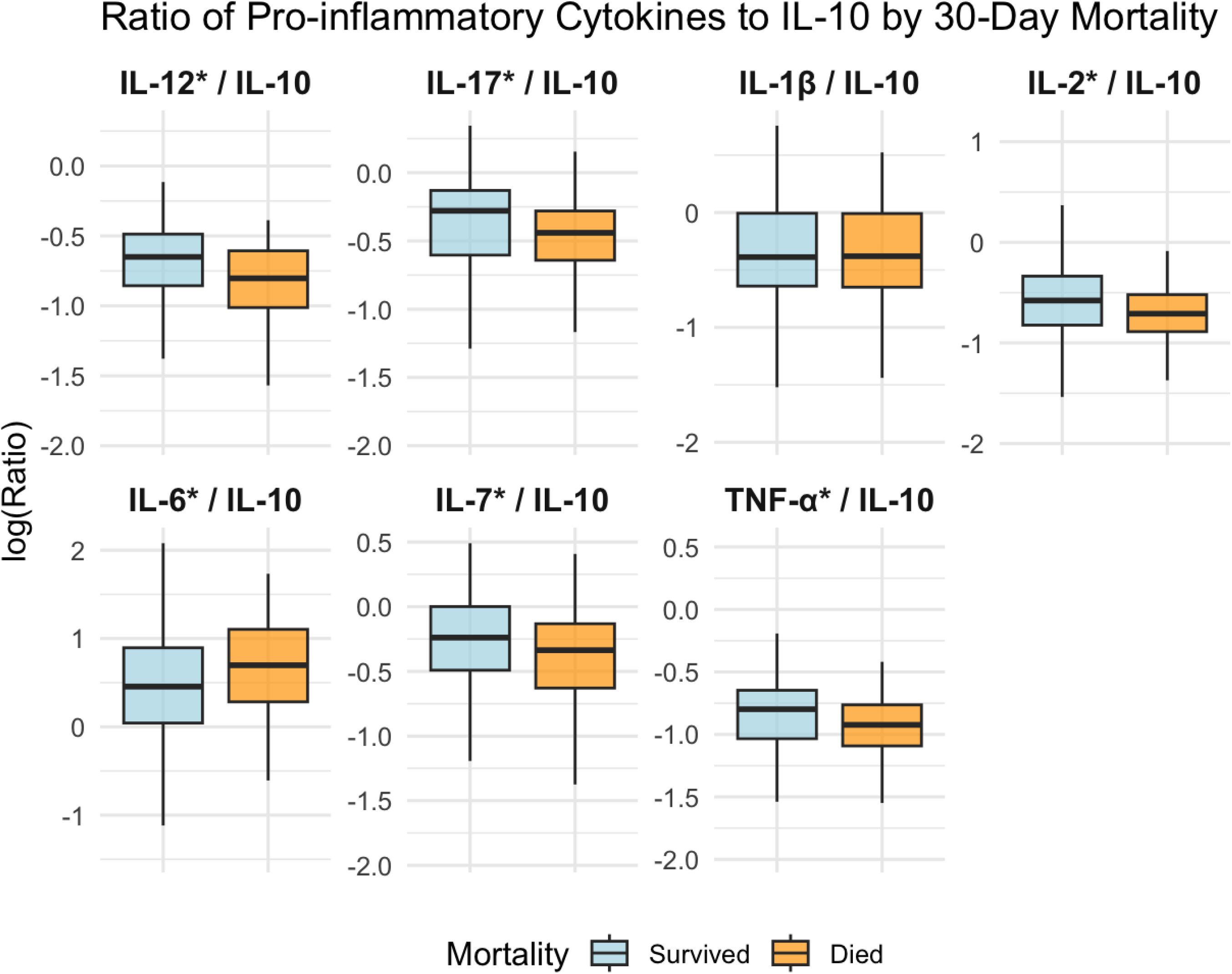
Box-and-whisker plots showing the median, interquartile range, and overall distribution of log-transformed ratios of pro-inflammatory cytokines to interleukin-10 among adults with sepsis in Uganda included in the discovery cohort, stratified by mortality.

**Supplementary figure 16.**
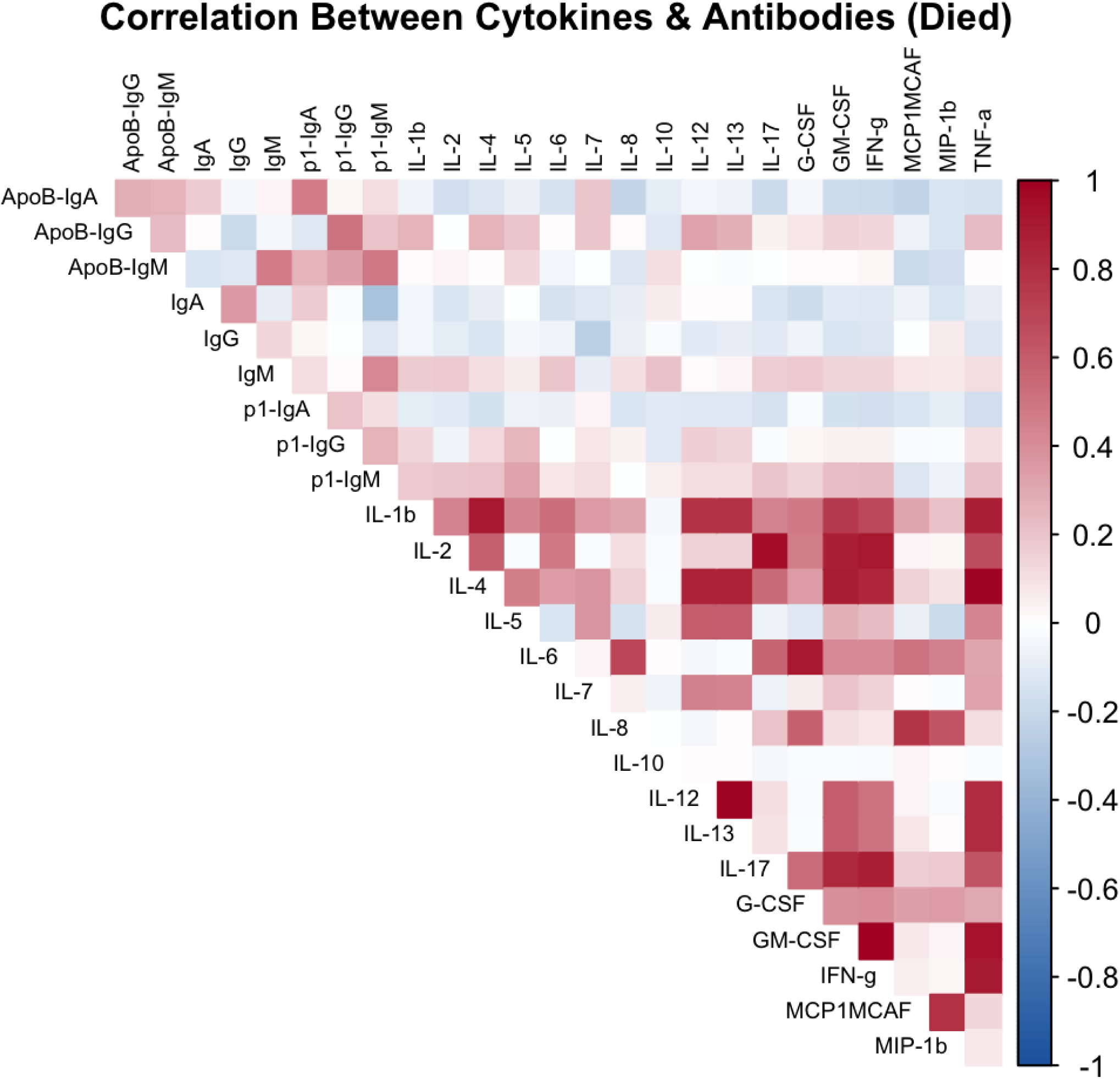
Correlation plot of cytokine and antibody concentrations among adults with sepsis in Uganda included in the discovery cohort who died. The heatmap values represent Pearson correlation coefficients between pairs of biomarkers.

## Notes

### Competing Interest Statement

The authors have declared no competing interest.

### Clinical Trial

NCT04618198

### Author Declarations

We obtained approval for the PRISM-U2 trial from ethics committees/institutional review boards of the Uganda National Council of Science and Technology (UNCST HS#419) and the University of Virginia (HSR# 13393). We obtained approval for the ATLAS trial from ethics committees/institutional review boards of the Tanzania Medicines and Medical Devices Authority (TMDA-WEB0021/CTR/0008/03), Uganda National Council of Science and Technology (UNCST HS1272ES) and the University of Virginia (HSR#200253) and registered at clinicaltrials.gov (NCT04618198). Participants or their next of kin provided informed consent prior to enrollment.

